# How immunity from and interaction with seasonal coronaviruses can shape SARS-CoV-2 epidemiology

**DOI:** 10.1101/2021.05.27.21257032

**Authors:** Naomi R Waterlow, Edwin van Leeuwen, Nicholas G. Davies, CMMID COVID-19 working group, Stefan Flasche, Rosalind M Eggo

**Affiliations:** Centre for Mathematical Modeling of Infectious Disease, London School of Hygiene and Tropical Medicine, UK; Statistics, Modelling and Economics Department, Public Health England, London, UK

**Author notes:** CMMID Working group: The following authors were part of the Centre for Mathematical Modelling of Infectious Disease COVID-19 Working Group. Each contributed in processing, cleaning and interpretation of data, interpreted findings, contributed to the manuscript, and approved the work for publication: Rachael Pung, Paul Mee, William Waites, Damien C Tully, Katherine E. Atkins, C Julian Villabona-Arenas, Graham Medley, Frank G Sandmann, Anna M Foss, Sophie R Meakin, Carl A B Pearson, Emilie Finch, Nikos I Bosse, Christopher I Jarvis, Kiesha Prem, Alicia Rosello, Kevin van Zandvoort, Rosanna C Barnard, Jiayao Lei, Yang Liu, Adam J Kucharski, Ciara V McCarthy, Sam Abbott, Emily S Nightingale, Joel Hellewell, Thibaut Jombart, David Hodgson, Gwenan M Knight, Amy Gimma, Yung-Wai Desmond Chan, Yalda Jafari, Samuel Clifford, Timothy W Russell, Fiona Yueqian Sun, Simon R Procter, Akira Endo, Oliver Brady, Kaja Abbas, Billy J Quilty, Mark Jit, Sebastian Funk, Fabienne Krauer, Matthew Quaife, Hamish P Gibbs, W John Edmunds, Mihaly Koltai, Kathleen O’Reilly, Rachel Lowe, James D Munday. The following funding sources are acknowledged as providing funding for the working group authors.This research was partly funded by the Bill & Melinda Gates Foundation (INV-001754: MQ; INV-003174: JYL, KP, MJ, YL; INV-016832: SRP; NTD Modelling Consortium OPP1184344: CABP, GFM; OPP1139859: BJQ; OPP1183986: ESN; OPP1191821: KO’R). BMGF (INV-016832; OPP1157270: KA). CADDE MR/S0195/1 & FAPESP 18/14389-0 (PM). EDCTP2 (RIA2020EF-2983-CSIGN: HPG). Elrha R2HC/UK FCDO/Wellcome Trust/This research was partly funded by the National Institute for Health Research (NIHR) using UK aid from the UK Government to support global health research. The views expressed in this publication are those of the author(s) and not necessarily those of the NIHR or the UK Department of Health and Social Care (KvZ). ERC Starting Grant (#757699: MQ). ERC.

**Keywords:** coronaviruses, immunity, SARS-CoV-2, COVID-19, cross-protection

## Abstract

We hypothesised that cross-protection from seasonal epidemics of human coronaviruses (HCoVs) could have affected SARS-CoV-2 transmission, including generating reduced susceptibility in children. To determine what the pre-pandemic distribution of immunity to HCoVs was, we fitted a mathematical model to 6 years of seasonal coronavirus surveillance data from England and Wales. We estimated a duration of immunity to seasonal HCoVs of 7.3 years (95%CI 6.8 - 7.9) and show that, while cross-protection between HCoV and SARS-CoV-2 may contribute to the age distribution, it is insufficient to explain the age pattern of SARS-CoV-2 infections in the first wave of the pandemic in England and Wales. Projections from our model illustrate how different strengths of cross-protection between circulating coronaviruses could determine the frequency and magnitude of SARS-CoV-2 epidemics over the coming decade, as well as the potential impact of cross-protection on future seasonal coronavirus transmission.

**Significance statement:** Cross-protection from seasonal epidemics of human coronaviruses (HCoVs) has been hypothesised to contribute to the relative sparing of children during the early phase of the pandemic. Testing this relies on understanding the pre-pandemic age-distribution of recent HCoV infections, but little is known about their dynamics. Using England and Wales as a case study, we use a transmission model to estimate the duration of immunity to seasonal coronaviruses, and show how cross-protection could have affected the age distribution of susceptibility during the first wave, and alter SARS-CoV-2 transmission patterns over the coming decade.

## Introduction

Due to the relatively short time since SARS-CoV-2 emerged, little is yet known about the duration of infection-induced immunity. While instances of confirmed reinfection of SARS-CoV-2 have been identified^1^, these are rare,^2^ indicating protection lasts for at least 6-8 months, which concurs with estimates from prospective studies^3,4^. For the success of long-term planning and design of control strategies against SARS-CoV-2, including vaccines, we must increase our understanding of the population-level immunity dynamics and interactions with other circulating viruses.

In order to evaluate the impacts of cross-immunity, we first need to quantify the duration of immune protection from seasonal coronaviruses. Four coronaviruses strains from two different genera are endemic in humans: two are alphacoronaviruses (HCoV-229E, HCoV-NL63) and two are betacoronaviruses (HCoV-HKU1, HCoV-OC43); SARS-CoV-2 is a member of the latter genera as are SARS-CoV-1 and MERS-CoV. In the UK, seasonal human coronavirus (HCoV) case incidence peaks January-February each year. The first infection with seasonal HCoVs typically occurs in childhood^5^ and reinfection with the same strain has been observed within a year^6,7^. However, there are also indications that immunity lasts longer, with few reinfections in a 3-year cohort study^8^ and sterilising immunity to homologous strains of HCoV-229E after one year in a challenge study^9^.

There may also be cross-protective immunity between seasonal HCoVs and SARS-family coronaviruses following infection. Human sera collected before the SARS-CoV-2 pandemic showed high IgG reactivity to seasonal HCoVs, but also low reactivity to SARS-CoV-2^10^, and SARS-CoV-1 infection induced antibody production against seasonal HCoVs^11^,12. Cross-reactive T-cells to SARS-CoV-2 have been found in 20%^13^ to 51%^14^ of unexposed individuals, with evidence that these responses stem from seasonal coronavirus infection^15^. It has also been noted that these are more prevalent in children and adolescents^16^.

Cross-protection from seasonal HCoVs may have, therefore, partially shaped the observed epidemiology of SARS-CoV-2. In particular, this cross-protection could help to explain the relatively low SARS-CoV-2 infection rate in children^17–21^. Since children likely have a higher annual attack rate of endemic HCoVs due to their higher contact rates^22^, they may be less susceptible to SARS-CoV-2 due to cross-protection. Using England and Wales as a case study, we use dynamic models to estimate: 1) the duration of infection-induced immunity to seasonal HCoVs, 2) the ability of potential cross-protection from seasonal HCoVs to explain the age patterns in the first wave of the SARS-CoV-2 pandemic, and 3) the implications of the duration of immunity and potential cross-protection on future dynamics of SARS-CoV-2.

## Results

### Seasonal HCoV and SARS-CoV-2 epidemic data

We extracted monthly, age group-stratified numbers of HCoV positive tests in England and Wales from the June 9, 2014 to February 17, 2020^23^ and daily number of COVID-19 deaths in England and Wales during the first wave of the pandemic (March 02, 2020 to June 01, 2020)^24^ (Figure 1). The timeframe for the HCoV data is from the first available date until February 2020 to avoid interference from SARS-CoV-2 transmission and reporting.

**Figure 1:**
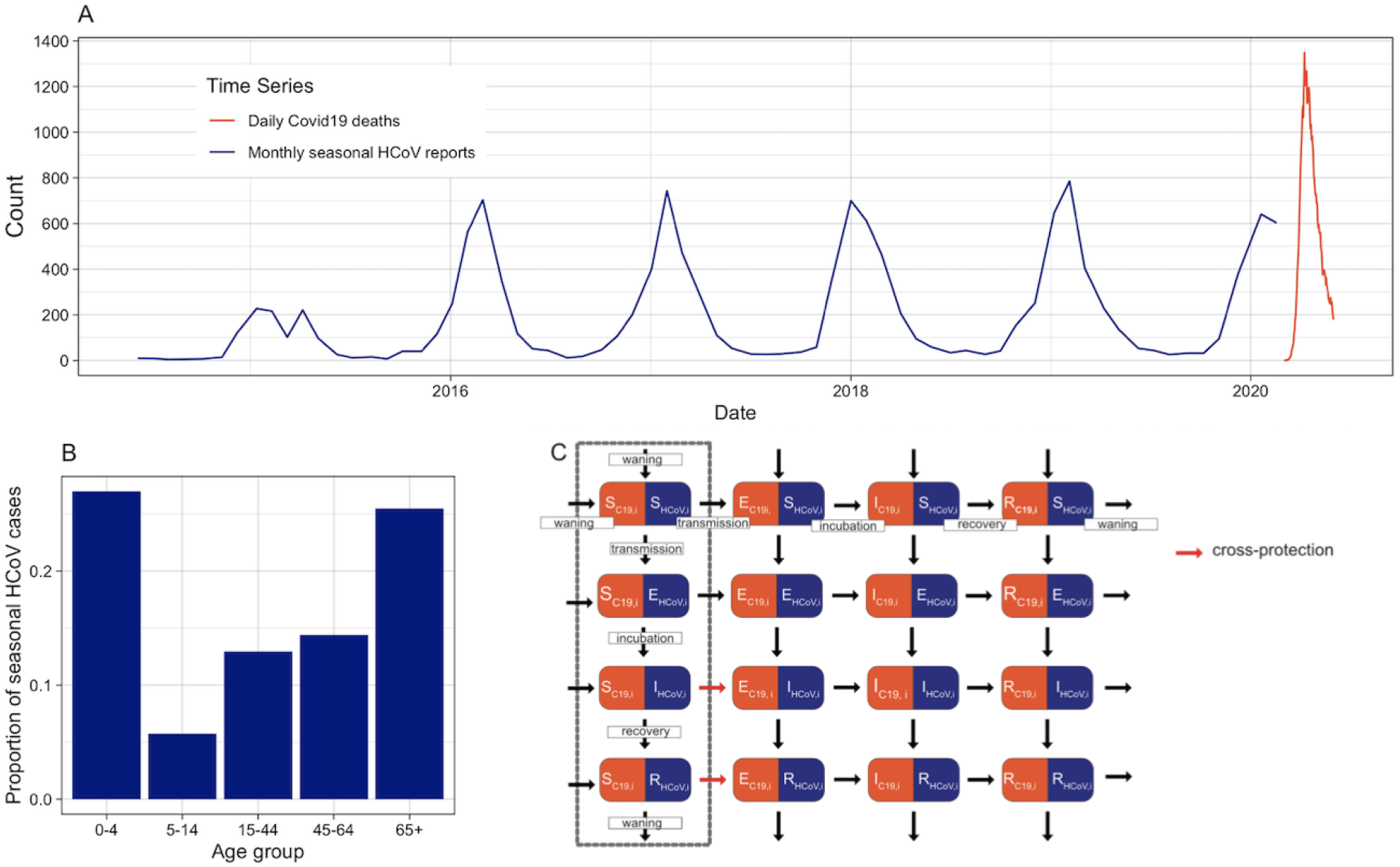
Data and model structure. HCoV is shown in blue and SARS-CoV-2 in red. A) Time series of reported HCoV cases and COVID-19 deaths. Left axis: reported seasonal HCoV cases to Public Health England (PHE), all ages. Right axis: COVID-19 daily deaths^24^. B) Proportion of seasonal HCoV cases reported by age group. C) Model diagram for seasonal HCoV (HCoV) and SARS-CoV-2 (C19). Individuals can be Susceptible (*S*), Exposed (*E*), Infectious (*I*) or Recovered (*R*) to SARS-CoV-2 or seasonal HCoVs. Following infection, individuals enter the exposed (*E*) category and then become infectious (*I*). They then recover (R) and via waning become fully susceptible again. At any point individuals can be infected by the other virus, although this is less likely to occur in the I and R categories, determined by the cross-protection parameter. For clarity, ageing, seasonality, births and deaths are not shown in this Figure. The initial model fit without SARS-CoV-2 only contains individuals within the compartments in the dashed gbox.

We fitted a dynamic transmission model using England and Wales as a case study (Figure 1C, box) using only the seasonal coronavirus model. Following infection, individuals are protected against infection with any seasonal HCoVs, with reinfection possible after a period of temporary but complete immunity. Waning of immunity against circulating viruses could result from decaying protection against homotypic viruses, and/or longer-lasting immunity against homotypic viruses but evolutionary change leading to immune escape ^25^. We do not track individual seasonal HCoV strains as available data are not sub-typed. We therefore assume that individual seasonal HCoV strain have the same parameter values, including duration of immunity and *R*_*0,HCoV*_. Transmission is seasonally forced using a cosine function.

### Immunity to seasonal HCoVs is estimated to last around 7 years

We fitted the model to the age group-specific seasonal HCoV data from June 09, 2014 until February 17, 2020, and estimated key seasonal HCoV parameters using parallel tempering^26^ (Figure 2). We fitted the duration of infection-induced immunity, the transmissibility, age-specific reporting proportions and two seasonal forcing parameters (Table S1). We estimated that the average duration of infection-induced immunity for seasonal HCoVs was 7.3 years (95% Credible Interval (CI): 6.8 - 7.9) and that the basic reproduction number was 5.7 (95% CI 5.4 - 6.0) (Figure 2B). Further details are given in the Supplement.

**Figure 2:**
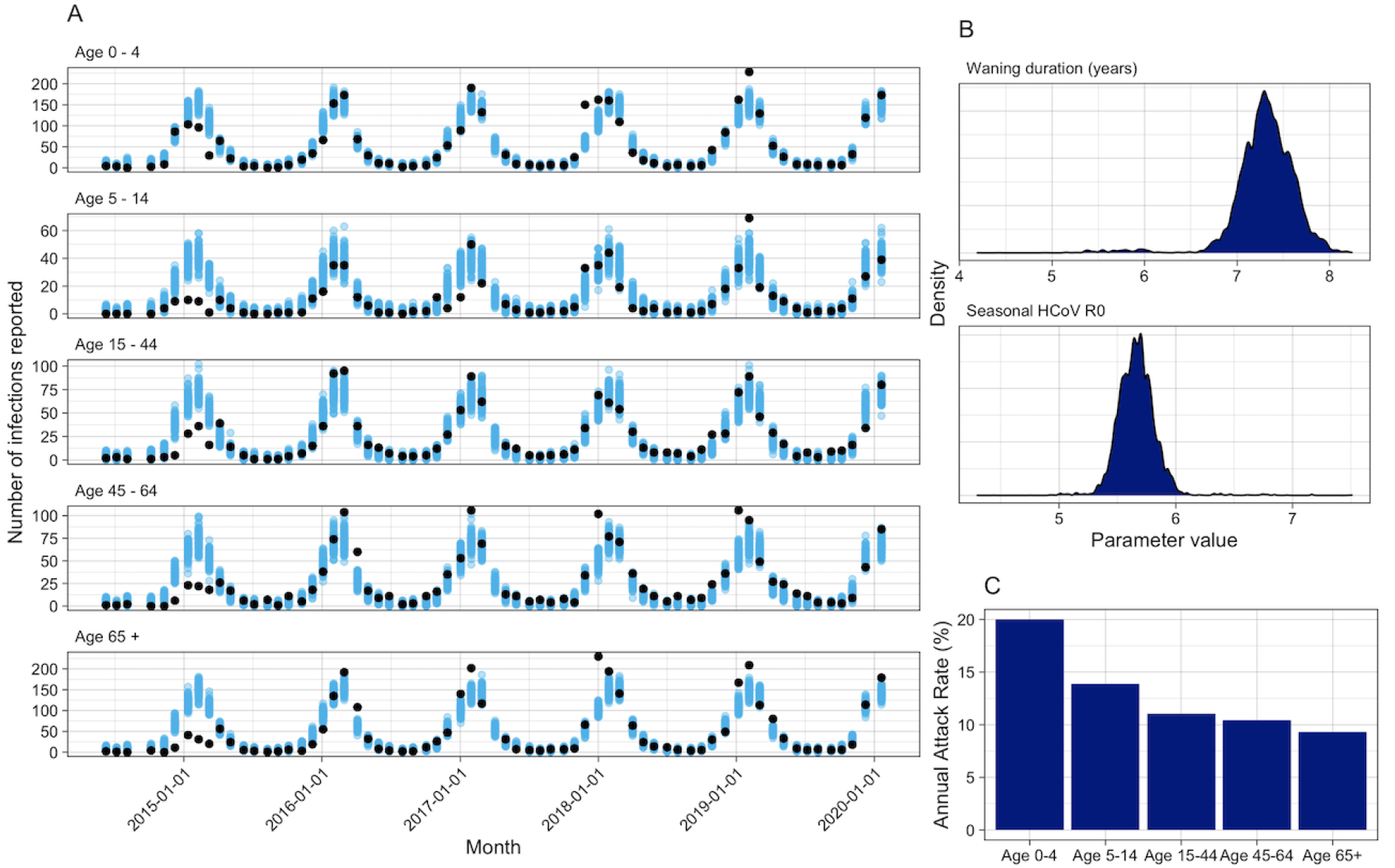
Seasonal HCoV Fit. A) Model fit for seasonal HCoV by age. Black dots show reported HCoV cases, blue are 100 random samples from the posterior. B) Posterior distributions for the duration of waning and the *R*_0_ of seasonal HCoV. C) Mean annual attack rate for each age group from 100 samples of the posterior and the last 5 years of the fit.

### Cross-protection from seasonal HCoVs is not sufficient to explain age-specific patterns of SARS-CoV-2 infection

We included SARS-CoV-2 into the model, where each compartment has the state for the combined seasonal HCoVs as well as the state for SARS-CoV-2 (Figure 1C, full model). We included cross-protection that decreases susceptibility to infection by SARS-CoV-2 by an amount, σ, for individuals in the *I*_*HCoV,i*_ or *R*_*HCoV,i*_ states (σ = 0 is no cross-protection and 1 is full cross-protection). We assume any interaction in the opposite direction would be negligible, due to the low proportion of the population that were infected in the first SARS-CoV-2 epidemic wave.

Using the posterior estimates of the seasonal HCoV parameters and the simulated output as initial states, we continued a simulation of epidemic seasonal HCoVs from the January 01, 2020 until June 01, 2020, including the introduction of SARS-CoV-2. Cross-protection from seasonal HCoVs and different mixing patterns (matching observed lockdown patterns, see Methods) were the only mechanisms we included that affected infection by SARS-CoV-2, so that we could evaluate the impact of cross-protection on the observed age distribution of cases.

For values of the cross-protection parameter between σ=0 and σ=1, we estimated *R*_*0,C19*_ and the number of introductions of SARS-CoV-2 by fitting the extended model to daily reported COVID-19 deaths (Figure 3a). We captured the national lockdown by decreasing contact rates following trends in Google mobility data^27^. Our model fits were able to closely match the reported mortality incidence for each value of the cross-protection parameter. However, the resulting *R*_*0,C19*_ varied widely, reaching over 25 for the strongest cross-protection (Figure 3B). The corresponding *R*_*eff,C19*_ before the intervention on March 23 ranged between 2.25 and 3.75 (see Supplement).

**Figure 3:**
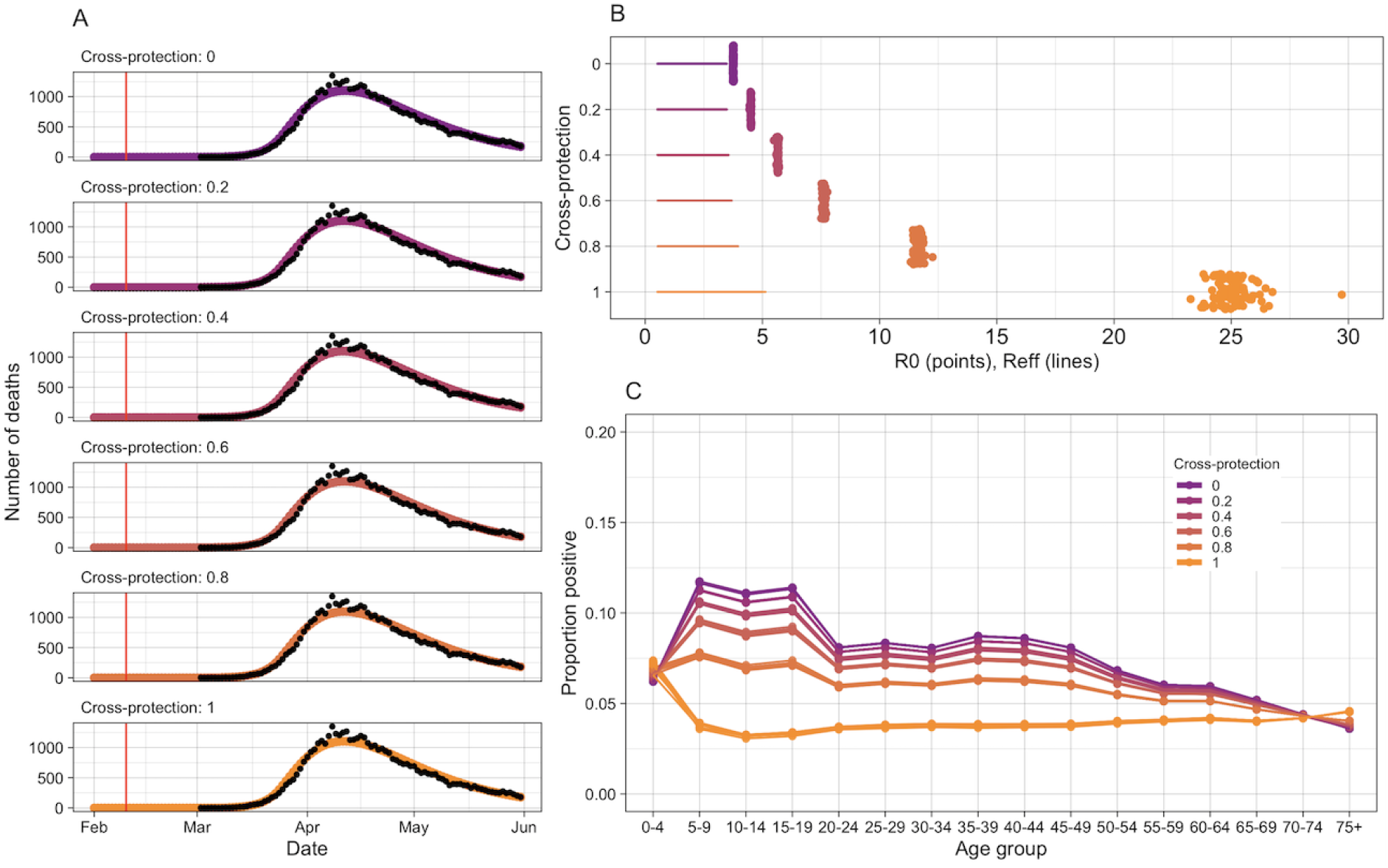
SARS-CoV-2 simulations. A) Fitted epidemic trajectories to daily SARS-CoV-2 deaths in 2020 in England and Wales^24^. Each facet is a different strength of cross-protection and contains output from 100 simulations. The red line indicates the fixed date when SARS-CoV-2 introduction starts. B) Estimated *R*_*0*_ values for SARS-CoV-2 with different strengths of cross-protection. Points display the *R*_*0, C19*_ and lines show the range of *R*_*eff, C19*_ during the simulation. C) Simulated age-specific serology rates for SARS-CoV-2 by the end of May 2020.

We then evaluated the age distribution of infections that would be detected by serology by the end of May in our model, across the range of values of the cross-protection parameter (Figure 1C). In simulations with no or low cross-protection the model predicted larger proportions of children to have been infected than in older age groups, differing from observed data^17,28^. As the strength of interaction increased, the age-distribution flattened and a smaller proportion of children became infected. With complete protection, there was a higher rate in the youngest age groups, which has not been observed^19–21,28^.

### Future SARS-CoV2 epidemiology could be shaped by coronavirus interactions

To determine possible long-term dynamics of interacting coronaviruses, we ran 30-year projections of our model including both HCoVs and SARS-CoV-2, with different assumptions on the strength of cross-protection and whether it acted from HCoV to SARS-CoV-2, or in both directions (Figure 4, and further samples in the supplement). In all scenarios we assumed no interventions, and used parameters estimated previously. For single-direction cross-protection, annual SARS-CoV-2 epidemics were projected to occur in scenarios with stronger cross-protection, whereas weaker / no cross-protection projected less frequent epidemics. However, strong cross-protection scenarios relied on very high and potentially unrealistic *R*_*0*_. In weaker cross-protection scenarios, interepidemic periods lasted multiple years following a pandemic. In scenarios with bi-directional cross-protection, SARS-CoV-2 infections also projected frequent epidemics, but led to the seasonal HCoV being disrupted. With low levels of cross-protection, SARS-CoV-2 and seasonal HCoV epidemics alternated, but as the cross-protection increased, SARS-CoV-2 epidemics became more frequent and outcompeted seasonal HCoV: while a cross-protection of 0.6 resulted in irregular dynamics of the viruses. At higher levels of cross-protection, no seasonal HCoV transmission occurred.

**Figure 4:**
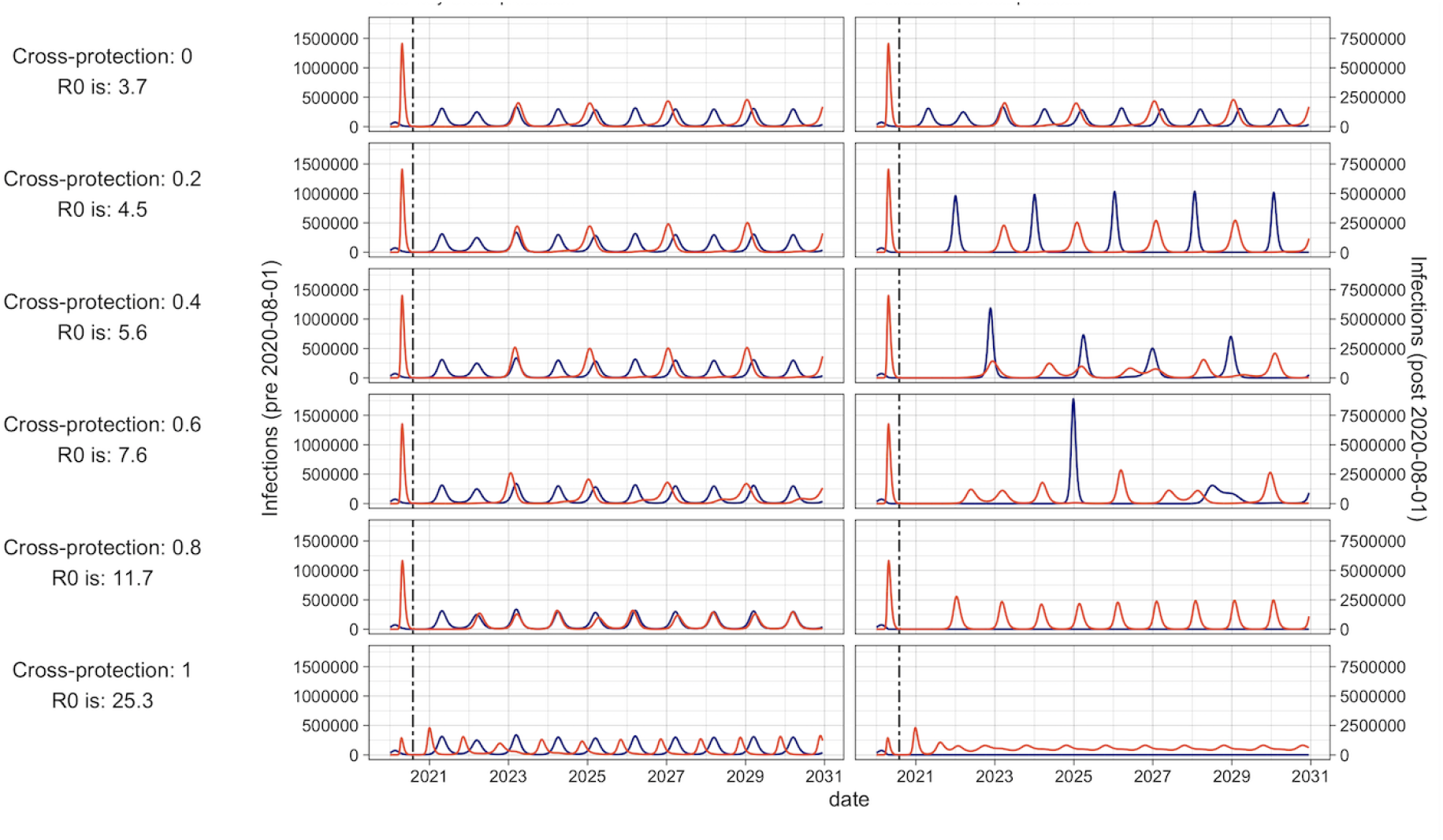
10-year forward projections of seasonal HCoV and SARS-CoV-2 epidemics. Red indicates SARS-CoV-2, blue indicates Seasonal HCoVs. The dashed vertical line indicates a change in axis scale due to the much larger SARS-CoV-2 pandemic wave, with that to the left of the dashed line marked by the left axis and that to the right by the right axis. Cross-protection strength and estimated SARS-CoV-2 *R*_*0*_ for the scenario are shown to the left of the figure. A has cross-protection from seasonal HCoV to SARS-CoV-2, and B has bidirectional cross-protection. No control measures were included.

## Discussion

We estimated that immunity to seasonal HCoVs lasts 7.3 years (95% CI: 6.8 - 7.9) and that the *R*_*0*_ is 5.7 (95% CI: 5.4 - 6.0). We found 12.8% (95% CI: 11.9 - 13.7%) reinfection within one year, and the median reinfection time was 5.1 years (95% CI: 4.7 - 5.5 years). It was possible to match the COVID-19 mortality data with the full range of cross-protection strengths between seasonal HCoV and SARS-CoV-2, but the estimated *R*_*0,C19*_ s were outside of a realistic range for very high values of cross protection, for example, a recent multi-setting study estimating the *R*_*0,C19*_ to be between 3.6 and 7.3^29^. Cross-protection from seasonal HCoVs to SARS-CoV-2 did not fully explain the apparent reduced susceptibility of children to SARS-CoV-2 observed during the first wave in the UK^17,18,20,21,28^. Future projections varied in the frequency of SARS-CoV-2 epidemics, with SARS-CoV-2 epidemics every two years at low levels of cross-protection, changing to annual epidemics with increased cross-protection. In scenarios with bi-directional cross-protection, epidemics were less predictable and SARS-CoV-2 out competed seasonal HCoVs. Further elucidating possible cross-protection and potential duration of protection is therefore critical for medium-to-long-term projections of SARS-CoV-2 epidemics.

Our estimates for the duration of homotypic protection following HCoV infection are comparable with other estimates, such as a cohort study where 8/216 (3%) confirmed infected individuals were reinfected over 5 years, and the median re-infection time in a study of 10 individuals^30^ varied between 30 and 55 months, depending on strain. However, estimates vary, with a larger study in Michigan estimating mean strain-specific reinfection to be between 19 and 33 months^31^, 19.9% of first infections being reinfected within 6 months in Kenya^32^, and a historical study, of just one seasonal HCoV strain (229E), estimating the time until T cells could no longer neutralise new strains at 8-17 years^25^. Other coronaviruses can also give indications on the duration of immunity, with T cells to SARS-CoV-1 detectable up to 11 years post-infection^33^. Using a similar modelling approach to that presented here, others have estimated that the average duration of immunity to seasonal coronavirus could be substantially shorter, under a year^34^. However, this implies very high annual attack rates, which are not observed in surveillance data, and our model suggests that a longer period of cross-protection may be more appropriate, and should be included in the proposed range of parameters for fitting such models.

Our model suggests that cross-protection between seasonal HCoVs and SARS-CoV-2 could account for some of the reduced susceptibility to infection of children in the first wave of the SARS-CoV-2 epidemic in England and Wales. Specifically, stronger cross-protection decreased the relative susceptibility to infection of children. This is in line with an American study showing that 50% of pre-pandemic donors had reactive T-cells to SARS-CoV-2^35^ and serological markers for a recent seasonal HCoV infection, suggesting that immune responses to seasonal HCoV could elicit cross-protective immunity. Moreover, 48% of uninfected individuals in a cohort from Australia had cross-reactive T-cells to SARS-CoV-2 and cross-reactivity was strongly correlated with memory of a seasonal coronavirus strains^15^. Other studies among healthy individuals without SARS-CoV-2 exposure found cross-reactive T-cells targeting SARS-CoV-2 in 51%^36^, 35%^14^ 24%^37^, and 20%^13^ of participants, suggesting a moderate amount of cross-immunity that likely stems from seasonal coronaviruses. There are indications that these cross-reactive T cells are present at higher frequency in younger vs older adults, correlating with our hypothesis that this could be due to increased infection from seasonal HCoVs^38^. Antibodies have also been shown to be cross-reactive^39^, although their persistence in the body is more varied and often shorter in duration than T-cells^40^. Cross-reactive responses have also been identified in other pandemic coronaviruses^10–12,41^, with some also showing cross-protection: SARS-CoV-1 and MERS-CoV T cell epitopes where protective in mice against other human and bat coronaviruses^42^. However a longitudinal study showed that whilst cross-reactive HCoV antibodies are boosted following SARS-CoV-2 infection, this does not correlate with protection against infection or hospitalisation^43^ and a lack of antibody-mediated neutralising cross-protection has been noted between sera from SARS-CoV-1 patients and SARS-CoV-2^44^. Therefore, whilst there is significant amounts of corroborating evidence that our find of some degree of cross-protection exists, the literature is not conclusive.

Our results indicate that cross-protection from seasonal coronaviruses alone cannot explain reduced susceptibility to infection of children. Other factors are needed to counteract the children’s higher than average exposure probability driven by their contact behavior^22^. One mechanism for this could be due to differences in children’s immune system^45^: children can produce broadly reactive antibodies that have not been influenced by commonly circulating pathogens and have different proportions of blood cell types, such as specific subtypes of memory B cells. Genetic analysis also suggests that cross-reactivity to SARS-CoV-2 antigens can not fully be explained by seasonal coronaviruses, implying that other unknown viruses/factors may induce cross-immunity^46^. We also modelled cross-protection as only reducing susceptibility to infection, whereas there could also be a reduction in transmission and/or disease severity^47–50^. Boosting of immunity by multiple infections has also been suggested to influence cross-protection^48^, where boosting by repeat infections was hypothesised to reduce the cross-protection to SARS-CoV-2. We did not include boosting in our model due to the added complexity.

The strength and implications of cross-protection between HCoVs and SARS-CoV-2 will become increasingly evident over the coming months and years. Our projections show that, depending on the extent of cross-protection, SARS-CoV-2 could eventually cause annual epidemics (strong cross-protection) or epidemics every 2 years (little cross-protection). If bi-directional cross-protection occurs, SARS-CoV-2 also has the ability to substantially disrupt seasonal HCoV transmission. This is based on our fit of the duration of immunity and the seasonal forcing parameters of seasonal HCoVs, which are likely to differ to some extent in the case of SARS-CoV-2. These scenarios are in line with others^34,51–53^, which suggest that ongoing SARS-CoV-2 transmission is likely. Our modelled projections assumed that no interventions were implemented. However, HCoV circulation was disrupted in winter 2020–2021^40^ likely due to social restrictions designed to curb the transmission of SARS-CoV-2. It is important to understand the longer-term dynamics of SARS-CoV-2, in order to minimise deaths and plan vaccination strategies. From an evolutionary perspective, cross-protection may be a strong driver for selection, so in the long run a less transmissible type with greater cross-protection against competing viruses may dominate.

We modelled all seasonal HCoVs as one virus, implicitly assuming complete cross-protection between them. There is evidence for cross-protection between seasonal HCoVs, such as the presence of cross-reactive antibodies^55^ and evidence from modelling studies^34^. Yet cross-protection may not be complete, or may be strain specific (alpha vs beta coronaviruses), and hence our assumption could lead to an underestimation of the true duration of protection. This is because the duration between subsequent homotypic infections would be longer than subsequent infections of any strain, for the same number of cases. We expect the assumption of modelling seasonal HCoVs to have a relatively small impact on the results of the cross-protection in the first wave of SARS-CoV-2, which uses the average cross-immunity profile at the end of the seasonal HCoV epidemic. However, the assumption may have a larger impact on the longer term dynamics, so these should be interpreted with caution. We also assumed that the strength of immunity to seasonal HCoVs is constant over repeated infections. An alternative mechanism would be that repeat infections strengthen immunity, as is hypothesised in some respiratory infections, such as Respiratory Syncytial Virus^56^, which could have led to a different estimate of immunity.

The emergence of SARS-CoV-2 has highlighted our lack of knowledge on coronavirus immunity and long term dynamics. In our study, we estimate that immunity against seasonal HCoVs can last years, however by necessity we made strong assumptions about the cross-immunity between seasonal HCoV strain. Further studies exploring cross-protection between strain for seasonal coronaviruses as well as routinely subtyped surveillance data would help inform future models. Nonetheless, based on the available data our study indicates that seasonal coronavirus immunity may last multiple years, which should be considered in the planning of subsequent studies. We also conclude that cross-protection from seasonal coronaviruses is not enough to explain the age susceptibility pattern of SARS-CoV-2, indicating other mechanisms must be involved. Whilst serological data could be useful to further evaluate the extent of cross-protection, the reduction in social contacts due to government interventions against SARS-CoV-2 complicates their use. Our models relies heavily on social contact matrices, and getting an accurate understanding of social contacts in the last year comes with many challenges, such as multiple changes in public health interventions with uncertain adherence. Our study highlights the importance of understanding the background environment of coronaviruses for insights into SARS-CoV-2 pandemic progression.

## Materials and Methods

We created a dynamic transmission model that includes cross-protection between seasonal HCoVs and SARS-CoV-2, using the UK as a case study (Figure 1). Initially, we fit the model without SARS-CoV-2 and estimated key seasonal HCoV parameters. Next we simulated SARS-CoV-2 introduction with varying strengths of cross-protection, to investigate the effect on age-specific susceptibility. The model was written in R^57^ and the code is available at https://github.com/cmmid/coronavirus_immunity

### Data

We extracted the monthly, age group-stratified number of HCoV positive tests in England and Wales between June 09, 2014 and February 17, 2020, from National Health Service (NHS) and Public Health England (PHE) laboratories^21^. The reported age groups are ages 0-4, 5-14, 15-44, 45-64 and 65+. We did not use data beyond February 2020, as we wanted to estimate seasonal HCoV parameters in the absence of SARS-CoV-2.

For SARS-CoV-2, we used the daily number of deaths with a confirmed SARS-CoV-2 positive test in the preceding 28 days from March 02, 2020 until May 31, 2020 reported by the Office for National Statistics (ONS)^24^.

### Cross-protection Model

We created a deterministic compartmental transmission model for coronavirus infections and their interactions. The population are either Susceptible (S), Exposed (E), Infectious (I) and Recovered (R) for both seasonal HCoVs and SARS-CoV-2. The subscripts used are “HCoV” for seasonal HCoVs and “C19” for SARS-CoV-2, with no differentiation between HCoV strains as the data are not sub-typed. Following infection, individuals enter the exposed category and become infected at rates λ_*HCoV,i*_ and λ_*C*19,*i*_ respectively, and individuals enter the infectious category at rates λ_HCoV_ and λ_C19._ They then recover and become fully susceptible again at rate ω. The force of infection for each virus is shown in equations 1 and 2. Each compartment in the model records the state for SARS-CoV-2 and seasonal HCoVs, with one for each combination of states (Figure 1) and all durations are exponentially distributed. At any point individuals can be infected by the other virus, although this is less likely to occur in the I and R categories, determined by the cross-protection parameter, σ. This takes into account both short term cross-protection from the activation of the immune system, and longer-term adaptive immunity. Both modelled viruses (HCoVs and SARS-CoV-2) are seasonally forced with a cosine function, which captures changes in seasonal human behaviour and climatic factors.

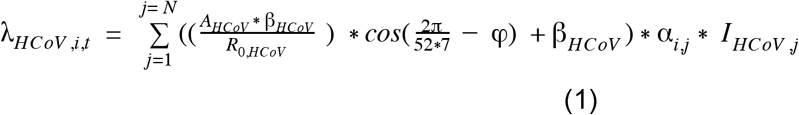

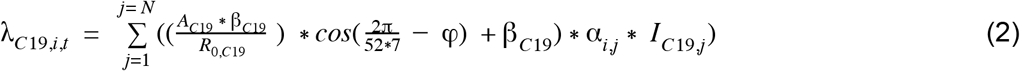

λ *= Force of infection, i,j = age groups, N = total number of age groups, A = seasonal amplitude, β = transmissibility, α = contact rates, I = number Infected, φ = timing of seasonal forcing*

As the seroprevalence for SARS-CoV-2 stayed below 5% during the modelled period, we assumed that the level of cross-protection conferred by SARS-CoV-2 on HCoV is negligible during the first epidemic wave. Cross-protection was the only mechanism we included for differing susceptibility to SARS-CoV-2 infection by age group, so that we could test whether it explained the observed infection pattern.

The modelled population was stratified into 5-year bands to 75+, with constant birth rates, matching death rates and ageing in line with the population of England and Wales (see Table 1).

**Table 1.**
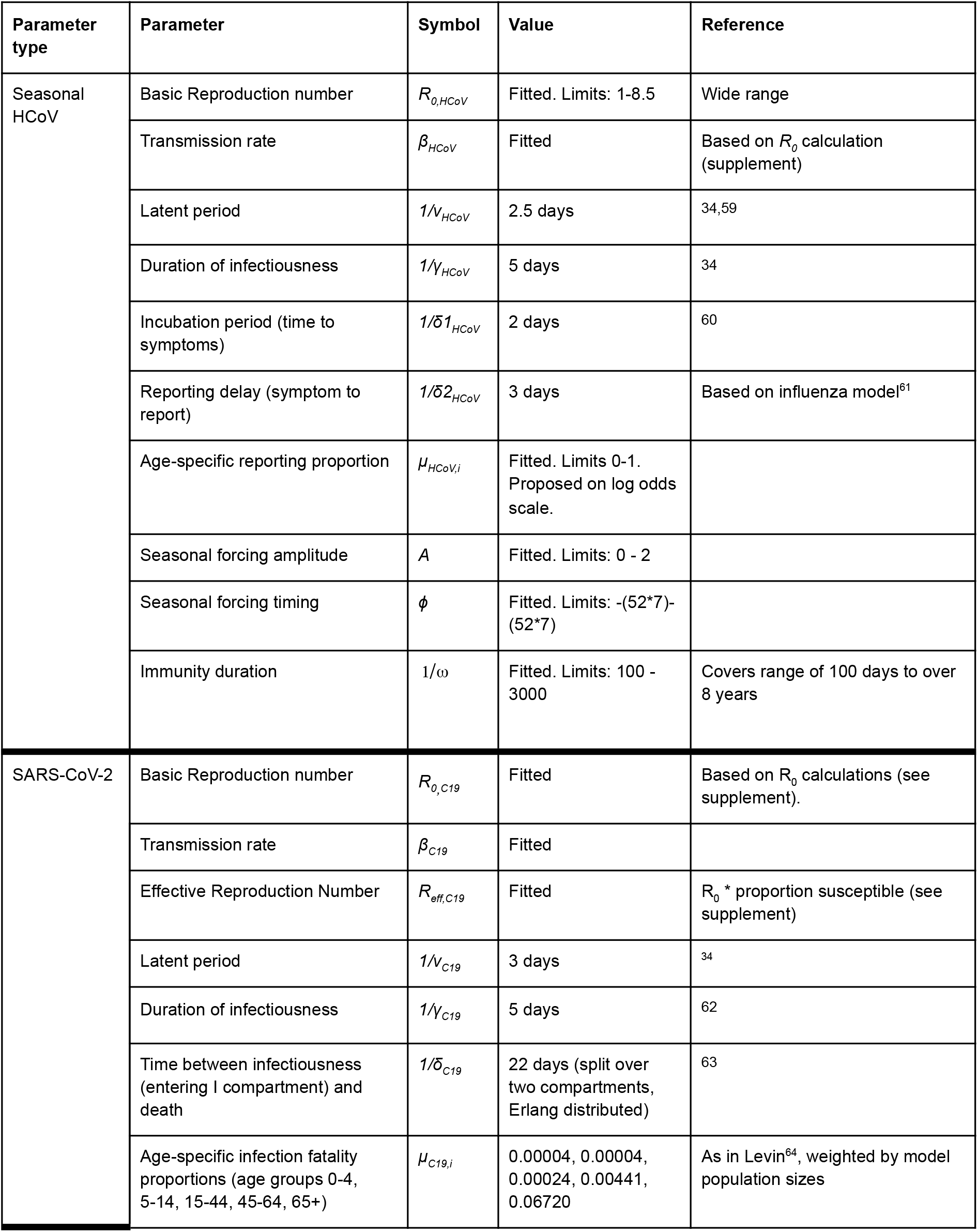
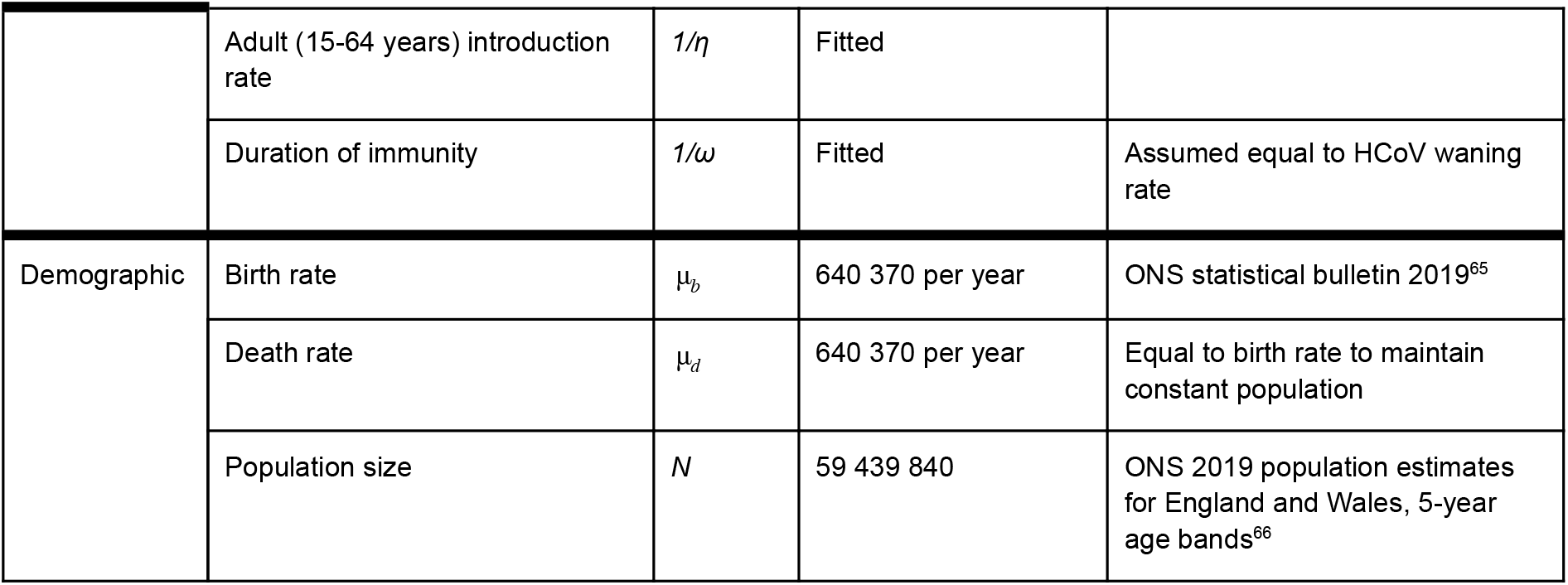
Model parameters.

Age-assortative mixing was modelled proportionately to patterns of conversational and physical contacts in the POLYMOD study^22^,58.

We ran a HCoV-only model for 15 years to reach equilibrium, and a further 5 years to generate simulations to match the data on seasonal HCoV cases from June 09, 2014 to February 17, 2020.

### Inferring seasonal HCoV parameters

We used reported seasonal HCoV cases from June 09, 2014 until February 17, 2020 to avoid overlap with SARS-CoV-2, where potential cross-protection could have occurred. We defined a binomial likelihood, where modelled infection incidence maps to reported cases via an age-dependent reporting proportion, *p*_*i*_. We assume equal reporting rates in age groups 5-15 and 45-65 to reduce the dimensions of the model, as initial fitting suggested these were very similar. The likelihood is therefore:

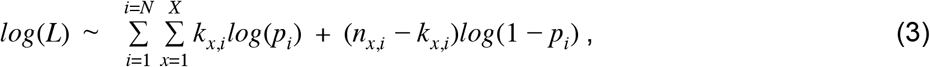

where *L* is the likelihood, *i* is age group to a total of N age groups, *x* are the reported monthly time points, *k*_*x,i*_ are the reported HCoV cases by age group, *n*_*x,i*_ are the model estimated infections per age group and *p*_*i*_ is the age-specific reporting rate.

We fit the model to the data using parallel tempering, adapted from Vousden et al.^22^ which is based on Monte Carlo Markov Chain (MCMC) inference. Unlike MCMC, multiple chains at different temperatures are run in parallel and swaps of parameter positions between chains are proposed. This allows more comprehensive exploration of the parameter space and allows the chains to move out of local maxima. We ran two sets of 16 chains and confirmed their convergence with the Gelman-Rubin statistic^67^, which was <1.1. We then combined the sample from both chains, excluding the burn-in, in order to increase sample size, resulting in 93900 samples. See supplement for more details.

The percentage infected within one year and the median duration to reinfection were calculated using distribution and quantile functions from the stats R package^57^.

### Simulating SARS-CoV-2 with a range of strengths of cross-protection

We drew 100 random samples from the joint posterior distribution and simulated daily deaths reported in the first wave of the SARS-CoV-2 epidemic in England and Wales, between March 02, 2020 and May 31, 2020. We explored the full range of possible cross-protection strengths, in each case fitting the transmission and introduction rates to the death data using maximum likelihood estimation with a Poisson likelihood. We therefore created 100 simulations of HCoV and SARS-CoV-2 circulation for each strength of cross-protection.

Due to the non-pharmaceutical interventions implemented in this period (“lockdown”), we adjust the contact matrices, which are split into three categories: school contacts, household contacts and all other contacts. From February 21, 2020, when Google mobility data becomes available, we adjust our “other” contacts in line with google mobility data. From February 23, 2020, we eliminate school contacts and assume that all remaining contacts are reduced to 33% of their transmission potential, due to social distancing and behavioral changes (“micro-distancing”)^68^. SARS-CoV-2 importations occur from February 15, 2020 until lockdown. See supplement for more details on the implementation of public health interventions.

To look at the proportion infected during the first wave we assumed that antibodies would take 3 weeks to rise to detectable levels after infection and not wane below the detection threshold during the study period^69^.

### Projecting future dynamics of SARS-CoV-2 and Seasonal HCoVs

We ran the model for 30 years, from January 01, 2020 without any changes in contacts, in order to project the future dynamics of SARS-CoV-2. As inputs, we used the estimated parameters from the seasonal HCoV fits, as well as the estimated transmission and introduction rates fitted for each of the samples. Projections for 10 of the samples are shown (in Figure 4 and Supplement).

## Data Availability

All data is previously published and referenced in the article: Respiratory infections: laboratory reports 2015-2020 - GOV.UK

## Classification

Medical Sciences, Applied Mathematics

## Notes

**Conflicts of interest:** None

**Funding:** NRW was supported by the Medical Research Council (grant number MR/N013638/1). EvL was supported by the National Institute for Health Research (NIHR) Health Protection Research Unit (HPRU) in Modelling and Health Economics, a partnership between PHE, Imperial College London, and LSHTM (grant number NIHR200908). EvL was supported by the European Union’s Horizon 2020 research and innovation programme - project EpiPose (101003688). SF is funded through a Sir Henry Dale Fellowship jointly funded by the Wellcome Trust and the Royal Society (grant number 208812/Z/17/Z). RME acknowledges an HDR UK Innovation Fellowship (grant: MR/S003975/1), MRC (grant: MC_PC 19065), and NIHR (grant: NIHR200908) for the Health Protection Research Unit in Modelling and Economics at LSHTM. The views expressed in this publication are those of the author(s) and not necessarily those of the NIHR or the UK Department of Health and Social Care.

### Competing Interest Statement

The authors have declared no competing interest.

### Funding Statement

NRW was supported by the Medical Research Council (grant number MR/N013638/1). EvL was supported by the National Institute for Health Research (NIHR) Health Protection Research Unit (HPRU) in Modelling and Health Economics, a partnership between PHE, Imperial College London, and LSHTM (grant number NIHR200908). EvL was supported by the European Union's Horizon 2020 research and innovation programme - project EpiPose (101003688). SF is funded through a Sir Henry Dale Fellowship jointly funded by the Wellcome Trust and the Royal Society (grant number 208812/Z/17/Z). RME acknowledges an HDR UK Innovation Fellowship (grant: MR/S003975/1), MRC (grant: MC_PC 19065), and NIHR (grant: NIHR200908) for the Health Protection Research Unit in Modelling and Economics at LSHTM. The views expressed in this publication are those of the author(s) and not necessarily those of the NIHR or the UK Department of Health and Social Care.
CMMID Working group: The following authors were part of the Centre for Mathematical Modelling of Infectious Disease COVID-19 Working Group. Each contributed in processing, cleaning and interpretation of data, interpreted findings, contributed to the manuscript, and approved the work for publication: Rachael Pung, Paul Mee, William Waites, Damien C Tully, Katherine E. Atkins, C Julian Villabona-Arenas, Graham Medley, Frank G Sandmann, Anna M Foss, Sophie R Meakin, Carl A B Pearson, Emilie Finch, Nikos I Bosse, Christopher I Jarvis, Kiesha Prem, Alicia Rosello, Kevin van Zandvoort, Rosanna C Barnard, Jiayao Lei, Yang Liu, Adam J Kucharski, Ciara V McCarthy, Sam Abbott, Emily S Nightingale, Joel Hellewell, Thibaut Jombart, David Hodgson, Gwenan M Knight, Amy Gimma, Yung-Wai Desmond Chan, Yalda Jafari, Samuel Clifford, Timothy W Russell, Fiona Yueqian Sun, Simon R Procter, Akira Endo, Oliver Brady, Kaja Abbas, Billy J Quilty, Mark Jit, Sebastian Funk, Fabienne Krauer, Matthew Quaife, Hamish P Gibbs, W John Edmunds, Mihaly Koltai, Kathleen O'Reilly, Rachel Lowe, James D Munday.
The following funding sources are acknowledged as providing funding for the working group authors. This research was partly funded by the Bill & Melinda Gates Foundation (INV-001754: MQ; INV-003174: JYL, KP, MJ, YL; INV-016832: SRP; NTD Modelling Consortium OPP1184344: CABP, GFM; OPP1139859: BJQ; OPP1183986: ESN; OPP1191821: KO'R). BMGF (INV-016832; OPP1157270: KA). CADDE MR/S0195/1 & FAPESP 18/14389-0 (PM). EDCTP2 (RIA2020EF-2983-CSIGN: HPG). Elrha R2HC/UK FCDO/Wellcome Trust/This research was partly funded by the National Institute for Health Research (NIHR) using UK aid from the UK Government to support global health research. The views expressed in this publication are those of the author(s) and not necessarily those of the NIHR or the UK Department of Health and Social Care (KvZ). ERC Starting Grant (#757699: MQ). ERC

### Author Declarations

LSHTM Research Ethics Committee

