## Supplemental Material for "How immunity from and interaction with seasonal coronaviruses can shape SARS-CoV-2 epidemiology"

1. Data **2**

2. Model equations **2**

3. R0 calculations **3**

4. Parallel Tempering **5**

5. Attack Rates **8**

6. Simulating lockdown **9**

7. Comparison with existing estimates **9**

8. Additional projections **10**

9. Seasonal sensitivity **16**

**References** **20**

#### 1. Data

We excluded one data point (April 03, 2017) as it was a duplicate of January 30, 2017, and due to the trend in the epidemic we assumed that January 30, 2017 was the correct one.

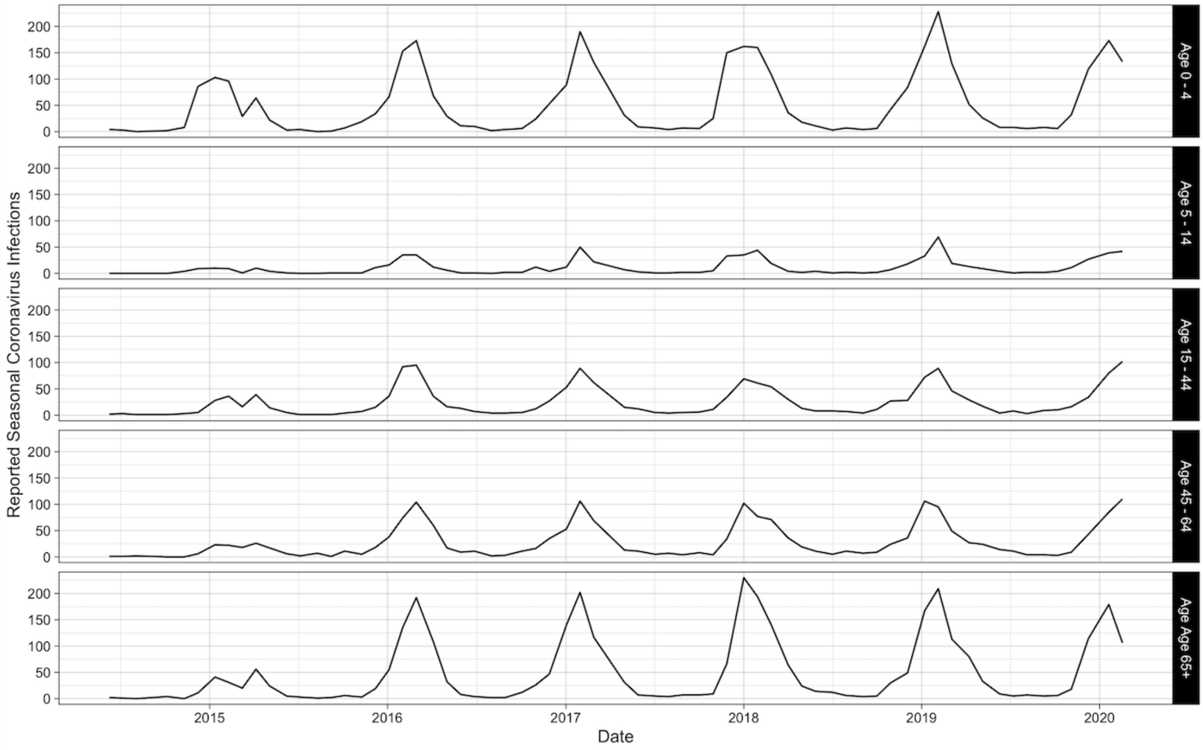


**Figure S1:Seasonal Coronavirus reported cases**

#### 2. Model equations

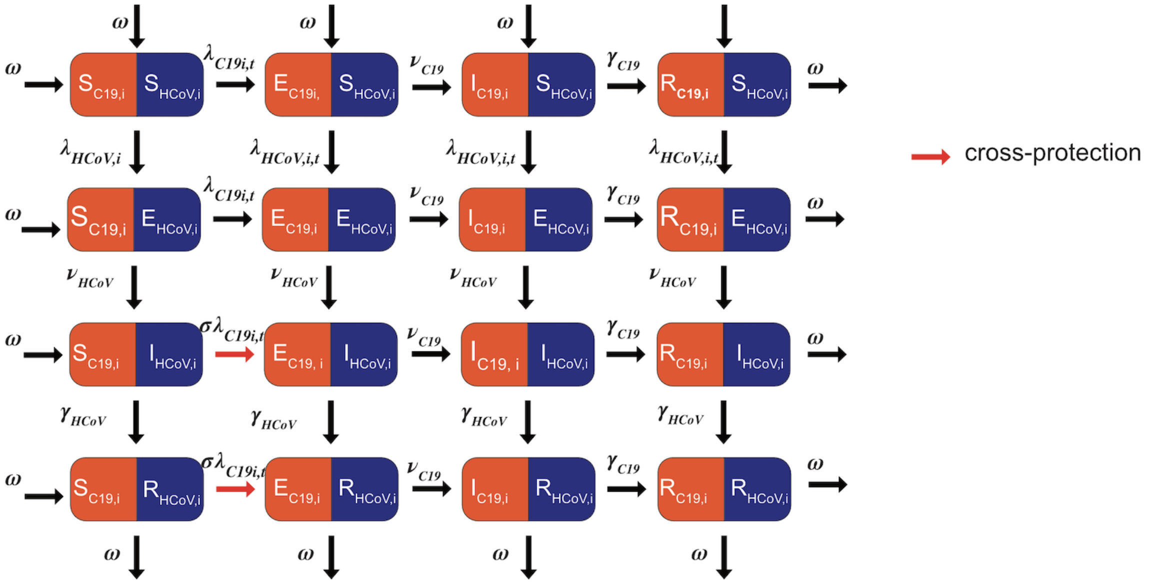


**Figure S1:Seasonal Coronavirus reported cases**. See below and table S1 for parameter definitions. For simplicity, only one age group is shown and ageing, births and deaths have not been included.

$$\lambda_{HCoV,i,t} = \sum_{j=1}^{j=N}((\frac{A_{HCoV} * \beta_{HCoV}}{R_{0,HCoV}}) * cos(\frac{2\pi}{52*7} - \varphi) + \beta_{HCoV}) * \alpha_{i,j} * I_{HCoV,j} \quad (1)$$

$$\lambda_{C19,i,t} = \sum_{j=1}^{j=N}((\frac{A_{C19} * \beta_{C19}}{R_{0,C19}}) * cos(\frac{2\pi}{52*7} - \varphi) + \beta_{C19}) * \alpha_{i,j} * I_{C19,j}) \quad (2)$$

$$\frac{dSS_i}{dt} = -\lambda_{C19,i} SS_i - \lambda_{HCoV,i,t}SS_i + \omega_{C19}RS_i + \omega_{HCoV}SR_i + \mu_{a,i}SS_{i-1} - \mu_{a,i+1}SS_i + \mu_{b,i} - \mu_{d,i}SS_i$$

$$\frac{dES_i}{dt} = -\nu_{C19}ES_i - \lambda_{HCoV,i,t}ES_i + \lambda_{C19,i} SS_i + \omega_{HCoV}ER_i + \mu_{a,i}ES_{i-1} - \mu_{a,i+1}ES_i - \mu_{d,i}ES_i$$

$$\frac{dIS_i}{dt} = -\gamma_{C19}IS_i - \lambda_{HCoV,i,t}IS_i + \nu_{C19}ES_i + \omega_{HCoV}IR_i + \mu_{a,i}IS_{i-1} - \mu_{a,i+1}IS_i - \mu_{d,i}IS_i$$

$$\frac{dRS_i}{dt} = -\omega_{C19}RS_i - \lambda_{HCoV,i,t}RS_i + \gamma_{C19}IS_i + \omega_{HCoV}RR_i + \mu_{a,i}RS_{i-1} - \mu_{a,i+1}RS_i - \mu_{d,i}RS_i$$

$$\frac{dSE_i}{dt} = -\lambda_{C19,i} SE_i - \nu_{HCoV}SE_i + \omega_{C19}RE_i + \lambda_{HCoV,i,t}SS_i + \mu_{a,i}SE_{i-1} - \mu_{a,i+1}SE_i - \mu_{d,i}SE_i$$

$$\frac{dEE_i}{dt} = -\nu_{C19}EE_i - \nu_{HCoV}EE_i + \lambda_{C19,i} SE_i + \lambda_{HCoV,i,t}ES_i + \mu_{a,i}EE_{i-1} - \mu_{a,i+1}EE_i - \mu_{d,i}EE_i$$

$$\frac{dIE_i}{dt} = -\gamma_{C19}IE_i - \nu_{HCoV}IE_i + \nu_{C19}EE_i + \lambda_{HCoV,i,t}RS_i + \mu_{a,i}IE_{i-1} - \mu_{a,i+1}IE_i - \mu_{d,i}IE_i$$

$$\frac{dRE_i}{dt} = -\omega_{C19}RE_i - \nu_{HCoV}RE_i + \gamma_{C19}IE_i + \lambda_{HCoV,i,t}RS_i + \mu_{a,i}RE_{i-1} - \mu_{a,i+1}RE_i - \mu_{d,i}RS_i$$

$$\frac{dSI_i}{dt} = -\sigma\lambda_{C19,i} SI_i - \gamma_{HCoV}SI_i + \omega_{C19}RI_i + \nu_{HCoV}SE_i + \mu_{a,i}SI_{i-1} - \mu_{a,i+1}SI_i - \mu_{d,i}SI_i$$

$$\frac{dEI_i}{dt} = -\nu_{C19}EI_i - \gamma_{HCoV}EI_i + \sigma\lambda_{C19,i} SI_i + \nu_{HCoV}EE_i + \mu_{a,i}EI_{i-1} - \mu_{a,i+1}EI_i - \mu_{d,i}EI_i$$

$$\frac{dII_i}{dt} = -\gamma_{C19}II_i - \gamma_{HCoV}II_i + \nu_{C19}EI_i + \nu_{HCoV}IE_i + \mu_{a,i}II_{i-1} - \mu_{a,i+1}II_i - \mu_{d,i}II_i$$

$$\frac{dRI_i}{dt} = -\omega_{C19}RI_i - \gamma_{HCoV}RI_i + \gamma_{C19}II_i + \nu_{HCoV}RE_i + \mu_{a,i}RI_{i-1} - \mu_{a,i+1}RI_i - \mu_{d,i}RI_i$$

$$\frac{dSR_i}{dt} = -\sigma\lambda_{C19,i} SR_i - \omega_{HCoV}SR_i + \omega_{C19}RR_i + \gamma_{HCoV}SI_i + \mu_{a,i}SR_{i-1} - \mu_{a,i+1}SR_i - \mu_{d,i}SR_i$$

$$\frac{dER_i}{dt} = -$$
$$\nu_{C19}ER_i - \omega_{HCoV}ER_i + \sigma\lambda_{C19,i} SR_i + \gamma_{HCoV}EI_i + \mu_{a,i}ER_{i-1} - \mu_{a,i+1}ER_i - \mu_{d,i}ER_i$$

$$\frac{dIR_i}{dt} = -\gamma_{C19}IR_i - \omega_{HCoV}IR_i + \nu_{HCoV}ER_i + \gamma_{HCoV}II_i + \mu_{a,i}IR_{i-1} - \mu_{a,i+1}IR_i - \mu_{d,i}IR_i$$

$$\frac{dRR_i}{dt} = -\omega_{C19}RR_i - \omega_{HCoV}RR_i + \gamma_{C19}IR_i + \gamma_{HCoV}RI_i + \mu_{a,i}RR_{i-1} - \mu_{a,i+1}RR_i - \mu_{d,i}RR_i$$

States
The first letter of the state indicates the state for SARS-CoV-2, the second letter indicates the state for HCoVs.
*S*: Susceptible
*E*: Exposed
*I*: Infected
*R*: Recovered

Subscripts
*C19*: SARS-CoV-2
*HCoV*: Seasonal HCoVs
*i, j:* age groups
*t*: time

| Symbol | Description | Value | Reference |
|---|---|---|---|
| $\lambda_{i,j}$ | Force of infection between age groups *i* and *j* | Estimated | Calculated as above |
| $\beta_{HCoV}$, $\beta_{C19}$ | Transmission rates | Estimated | |
| $\alpha_{i,j}$ | Contact rate between age groups *i* and *j* | Fixed | POLYMOD[1] |
| *N* | Total number of age groups | 16 | 5 year age-bands, up to 75+ |
| *A* | Seasonal amplitude | Estimated | |
| $\varphi$ | Timing of seasonal forcing | Estimated | |
| $\sigma$ | 1 - strength of cross-protection | Estimated | |
| $v_{HCoV}$ | Seasonal HCoV rate of latency | 1/2.5 days | [2] |
| $v_{C19}$ | SARS-CoV-2 rate of latency | 1/3 day | [3] |
| $\gamma_{HCoV}$ | Seasonal HCoV rate of recovery | 1/5 days | [3] |
| $\gamma_{C19}$ | SARS-CoV-2 rate of recovery | 1/5 days | [4] |
| $\omega$ | Rate of loss of immunity | Estimated | |
| $\mu_{a,i}$ | Ageing rate | Fixed | 1/length of age group |
| $\mu_{b,i}$ | Annual birth rate ( 0 unless i == 1) | 1/ 640 370 | ONS statistical bulletin 2019[5] |
| $\mu_{d,i}$ | Annual death rate | 1/ 640 370 | Equal to birth rate, to maintain constant population |

**Table S1 : Model parameters**

### 3. $R_0$ calculations

We used the method described by Diekmann *et al.* (2009)[6] to calculate the $R_0$ for each virus. The dominant eigenvalue of the matrix is the $R_0$ of the matrix $-T\Sigma^{-1}$, where $T$ is the transmission part of the Jacobian matrix, describing new infections and $\Sigma$ is the transition part, describing changes in the infectious state. See reference for further details. For seasonal HCoVs we used the base transmission rate.

For each age group, compartments in the matrix are SE and SI, and ES and IS, as we calculate the $R_0$ assuming no cross-protection. The first row/column represents the E compartments, and the second row represents the I compartment for the first age group. Only the transmissions for the first age group are shown. Three dots (…) represent the pattern continuing, one dot (.) represents equations not shown because they do not refer to the first age group.

$$\mathrm{T}_{C19} = \begin{vmatrix} 0 & \beta_{C19}\,\alpha_{i,j} & 0 & \beta_{C19}\,\alpha_{i,j} & \dots & \beta_{C19}\,\alpha_{i,j} \\ 0 & 0 & 0 & 0 & \dots & 0 \\ 0 & \beta_{C19}\,\alpha_{i,j} & . & . & . & . \\ 0 & 0 & . & . & . & . \\ 0 & \beta_{C19}\,\alpha_{i,j} & . & . & . & . \\ \dots & \dots & . & . & . & . \\ 0 & \beta_{C19}\,\alpha_{i,j} & . & . & . & . \end{vmatrix} \qquad \Sigma_{C19} = \begin{vmatrix} -\nu_{C19} - \mu\alpha_i & 0 & 0 & \dots & 0 \\ -\nu_{C19} & -\gamma_{C19} - \mu\alpha_i & 0 & \dots & 0 \\ \mu\alpha_{i+1} & 0 & . & . & . \\ 0 & \mu\alpha_{i+1} & . & . & . \\ 0 & 0 & . & . & . \\ \dots & \dots & . & . & . \\ 0 & 0 & . & . & . \end{vmatrix}$$

$$\mathrm{T}_{HCoV} = \begin{vmatrix} 0 & \beta_{HCoV}\,\alpha_{i,j} & 0 & \beta_{HCoV}\,\alpha_{i,j} & \dots & \beta_{HCoV}\,\alpha_{i,j} \\ 0 & 0 & 0 & 0 & \dots & 0 \\ 0 & \beta_{HCoV}\,\alpha_{i,j} & . & . & . & . \\ 0 & 0 & . & . & . & . \\ 0 & \beta_{HCoV}\,\alpha_{i,j} & . & . & . & . \\ \dots & \dots & . & . & . & . \\ 0 & \beta_{HCoV}\,\alpha_{i,j} & . & . & . & . \end{vmatrix} \qquad \Sigma_{HCoV} = \begin{vmatrix} -\nu_{HCoV} - \mu\alpha_i & 0 & 0 & \dots & 0 \\ -\nu_{HCoV} & -\gamma_{HCoV} - \mu\alpha_i & 0 & \dots & 0 \\ \mu\alpha_{i+1} & 0 & . & . & . \\ 0 & \mu\alpha_{i+1} & . & . & . \\ 0 & 0 & . & . & . \\ \dots & \dots & . & . & . \\ 0 & 0 & . & . & . \end{vmatrix}$$

For the SARS-CoV-2 simulations we calculated the $R_{eff}$ through time (Figure S1), which is influenced by the $R_0$, the level of cross-protection and the seasonal HCoV circulation. The $R_{eff}$ is the largest eigenvalue of the NGM where each row is multiplied by the proportion susceptible in that age group. Estimates for the $R_{effective}$ in the UK of SARS-CoV-2 before lockdown were between 2.25 and 3.75, so we used these as boundaries. During this period it was only the highest level of cross-protection that did not have an appropriate $R_0$.

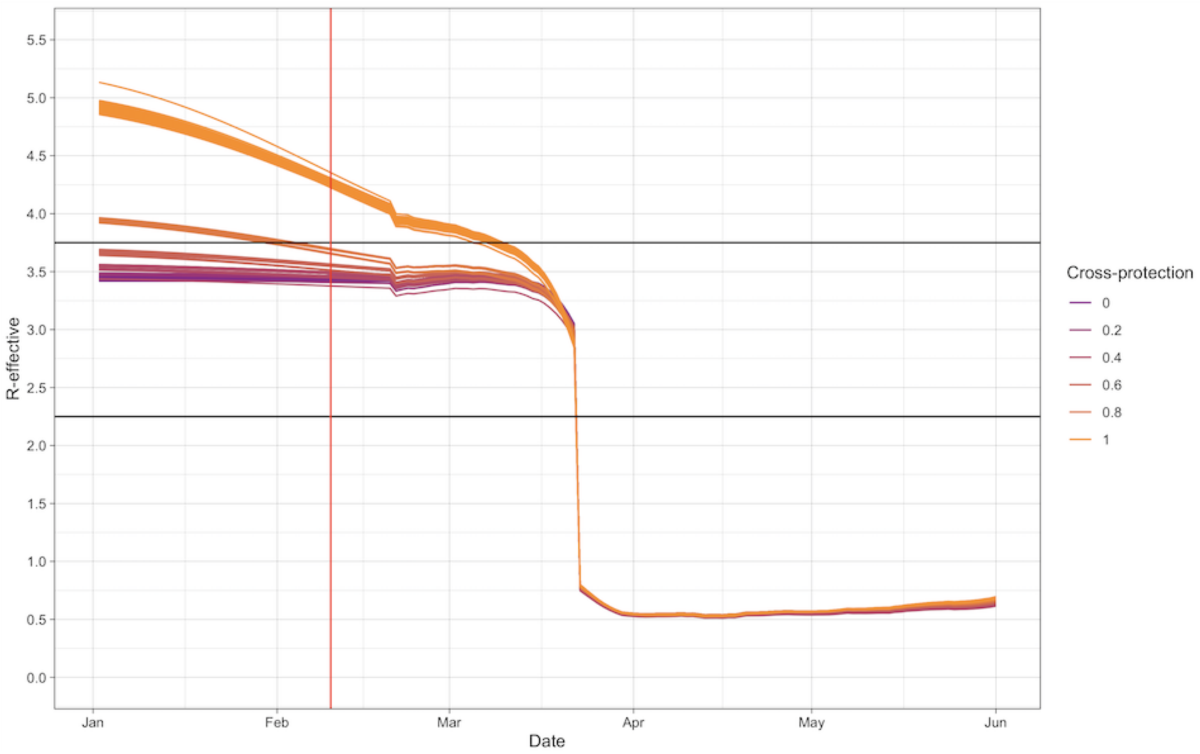


**Figure S3: $R_{effective}$ values over time for SARS-CoV-2.** Blue lines indicate $R_{effective}$ for simulations at different levels of cross-protection. Black lines show the $R_{effective}$ limits of 2.25 and 3.75 and the red line shows the date of SARS-CoV-2 introduction.

#### 4. Parallel Tempering

We proposed chains to swap with the chain of the next lowest temperature every 5 iterations of the MCMC. The highest and lowest temperatures were fixed at 1000 and 1 (the null chain). The number of chains was then adjusted to achieve an acceptance rate of swaps of between 0.15 and 0.25 . Swaps were accepted based on our swapping equation, adapted from Vousden *et al*[7] following the equation:

$$R = \frac{e^{(LL(i) - LL(j))}}{\tau_j - \tau_i}$$

Where

$$\tau_i = \frac{1}{T_i}$$

And $T_i$ is the temperature of chain i, and LL(i) is the log likelihood of chain i.

We then ran the parallel tempering algorithm from multiple different start values, each with 16 chains. Within each chain parameters were proposed using a covariance matrix. This resulted in two converged chains, which we confirmed by checking that the Gelman-Rubin statistic[8] was <1.1. We discarded 12000 iterations of each as burn in and then combined the samples from the two converged regions to increase the sample size. A set of chains is shown in Figure S2. Figure S3 shows the posterior distribution for all parameters and they are summarised in Table S1.

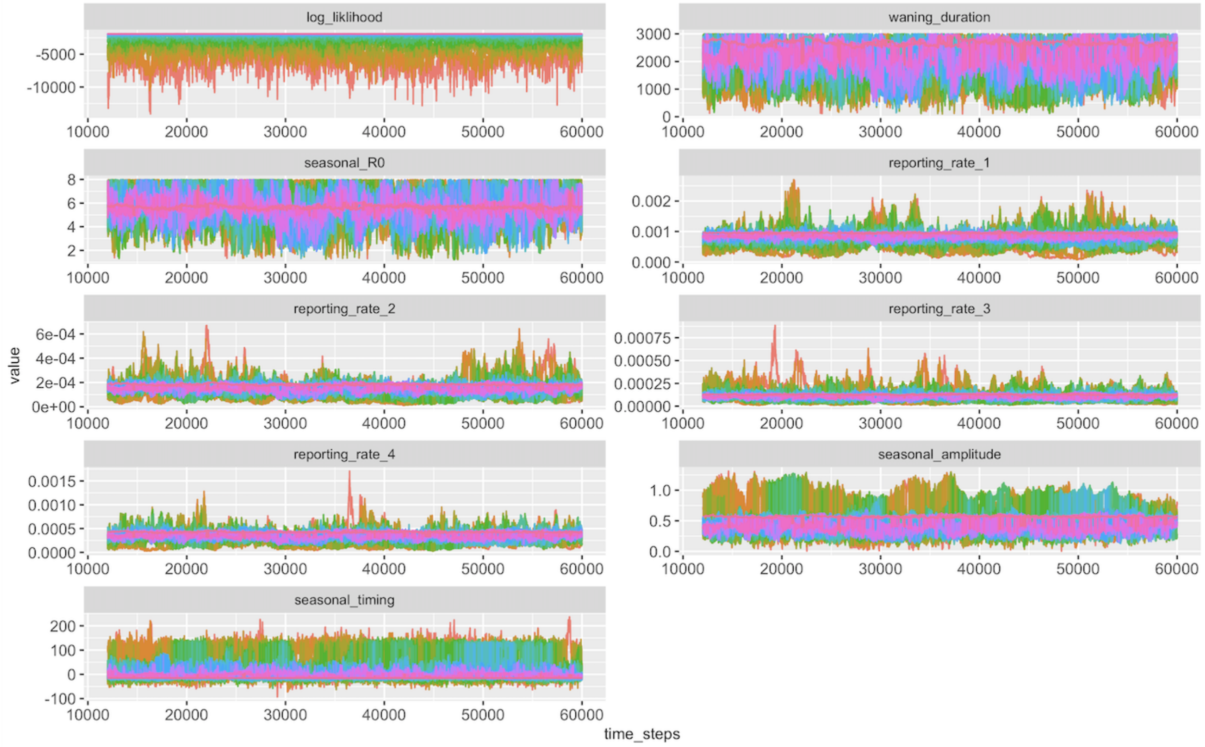


**Figure S4: Trace plots of one replicate showing all 16 chains.** Each colour is one chain, with bright pink being the coolest (null) chain.

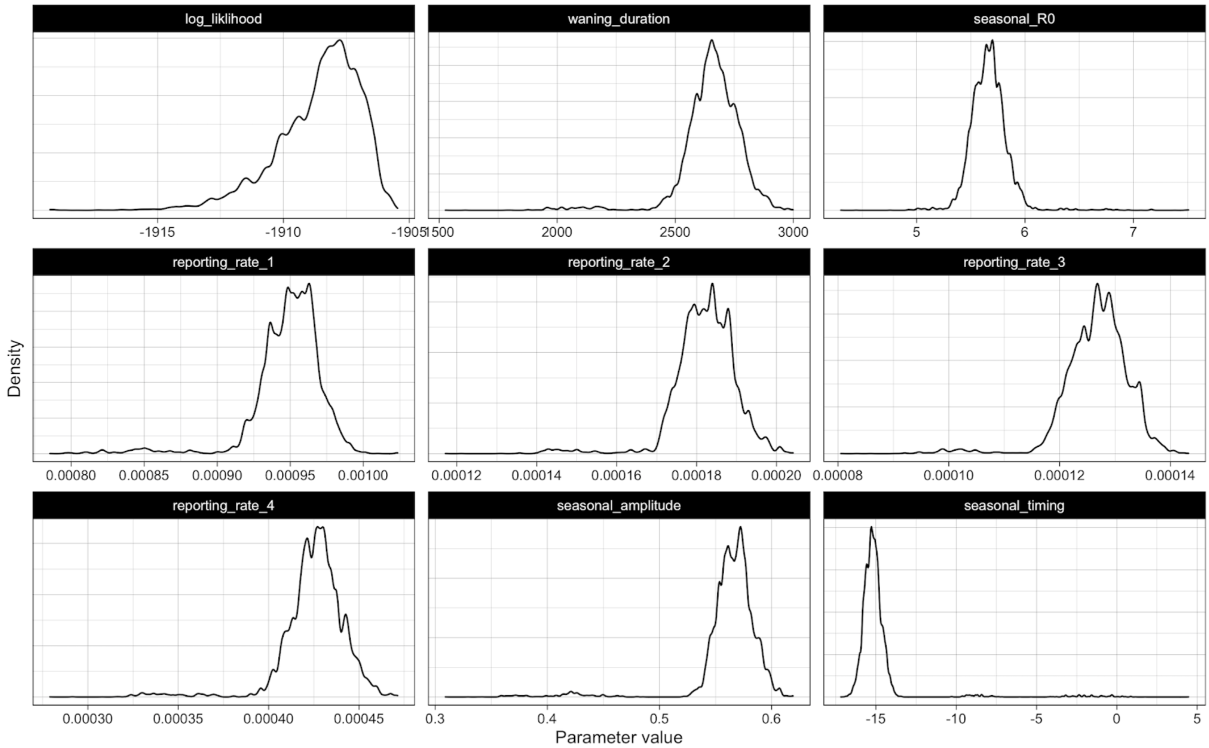


**Figure S5: Posterior distributions of fitted parameters for the HCoV fit.**

| Parameter | Symbol | Median (95% CI) |
|---|---|---|
| Basic Reproduction number | $R_{0,HCoV}$ | 5.7 (5.4 - 6.0) |
| Immunity duration | $\omega$ | 7.3 (6.8 - 7.9) |
| Age-specific reporting proportion 0-4 | $\mu_{HCoV,i}$ | 0.00095 (0.00092 - 0.00099) |
| Age-specific reporting proportion 5 - 14, 45-64 | $\mu_{HCoV,i}$ | 0.00018 (0.00017 - 0.00020) |
| Age-specific reporting proportion 15-44 | $\mu_{HCoV,i}$ | 0.00013 (0.00012 - 0.00014) |
| Age-specific reporting proportion 65+ | $\mu_{HCoV,i}$ | 0.00043 (0.00040 - 0.00045) |
| Seasonal forcing amplitude | A | 0.57 (0.54 - 0.60) |
| Seasonal forcing timing | $\phi$ | -15.2 (-16.4 - -14.0) |

**Table S2: Median and 95% quantiles of the posterior distributions of fitted HCoV parameters.**

Figure S2 shows a heatmap of the likelihood of the model at different value of $R_0$ and durations of immunity waning. One sample from the posterior of the fit was taken, and all parameter values apart from the $R_0$ and the duration of waning were kept constant. The log likelihood was then calculated for each combination of $R_0$ and waning durations. The fitted value is shown with a +.

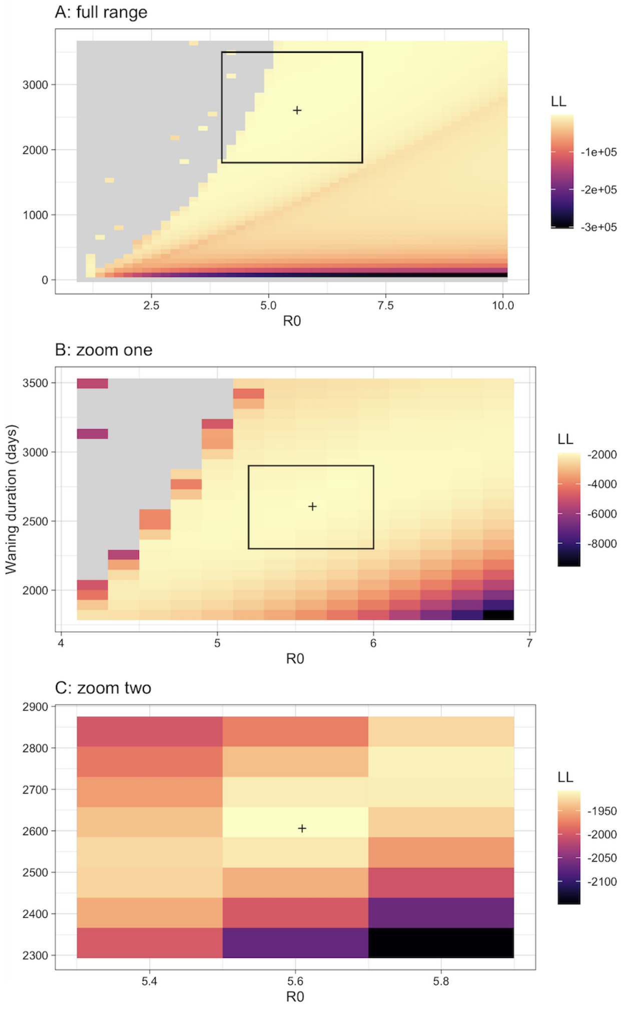


**Figure S6: Likelihood plane with varying $R_0$ and duration of waning**. A) shows the full range B) and C) show zoomed in areas. Colour indicates the likelihood value. The black rectangles show the area zoomed in on and the '+' symbol shows the estimated values from the parallel tempering fits.

### 5. Attack Rates

Average annual attack rates for the seasonal HCoV are shown in Table S2. This is the mean attack rate for each age group, averaged over 100 samples from the joint posterior and the last 5 years in our seasonal HCoV fit.

| **Age group** | **Attack rate (%)** |
|---|---|
| < 5 | 19.9 |
| 5 - 14 | 13.9 |
| 15 - 44 | 11.0 |
| 45 - 64 | 10.4 |
| 65+ | 9.3 |

**Table S2: Mean attack rates by age group.** Values given are mean across 100 random samples from the posterior and the last 5 years of the seasonal HCoV fit.

### 6. Simulating lockdown

Due to the non-pharmaceutical interventions implemented in this period (“lockdown”), we adjust the contact matrices, which are split into three categories: school contacts, household contacts and other contacts. Other contacts included all other categories reported in the polymod dataset. We then adjusted the contacts as follows.

- From February 21, 2020 (when google mobility data first becomes available), we adjust our ‘other’ contacts group by the average change in retail/recreation, workplace, grocery/pharmacy and transit stations, according to the Google Mobility UK records
- From March 23, 2020 (lockdown including school closures), school contacts are reduced to 0 with no re-attribution of those contacts.
- From March 23, 2020 (lockdown), Other and household contacts are multiplied by a ‘social distancing factor’ which we set at 0.33. This simulates other interventions such as social distancing, increased hand washing and mask wearing, and we chose the value based on being within a plausible range and appropriate looking simulations. We adapt households as the original contact matrix includes all household interactions, including visitors.
- Importations occur from the date of SARS-CoV-2 introduction (February 15, 2020) in the UK until the March 23, 2020 (lockdown). The date of SARS-CoV-2 introduction was chosen as a plausible value that allowed the simulated deaths to peak at the right time of year.

### 7. Comparison with existing estimates

As our estimate of the duration of immunity is longer than other estimates, we compared it to parameters estimated in the 2020 Kissler et al. paper[3]. While the states are the same in the two model, there are some key differences including:

- Kissler et al. only model beta-coronaviruses, whereas we model all seasonal coronaviruses
- Kissler et al. model the two coronaviruses separately and estimate the cross-protection between them. We are instead modelling all coronaviruses together, thereby implicitly assuming complete cross-protection.
- The Kissler et al. model is not age structured
- The latency period in the Kissler et al. model is slightly longer (3 days instead of 2 days).

We investigated the impact of using their estimated parameter values in our model. We fixed their estimated values of $R_0$ (2), waning (45 weeks) and seasonality parameters, as well as changing our model to match their longer latency period. We then fit the reporting rates using Maximum Likelihood Estimation (L-BFGS-B optimisation) to fit the seasonal coronavirus, using the same likelihood as in the main paper. While the model still produced what looked like a good fit (See Fig S4), the log likelihood values were significantly lower (-2235 vs -1905).

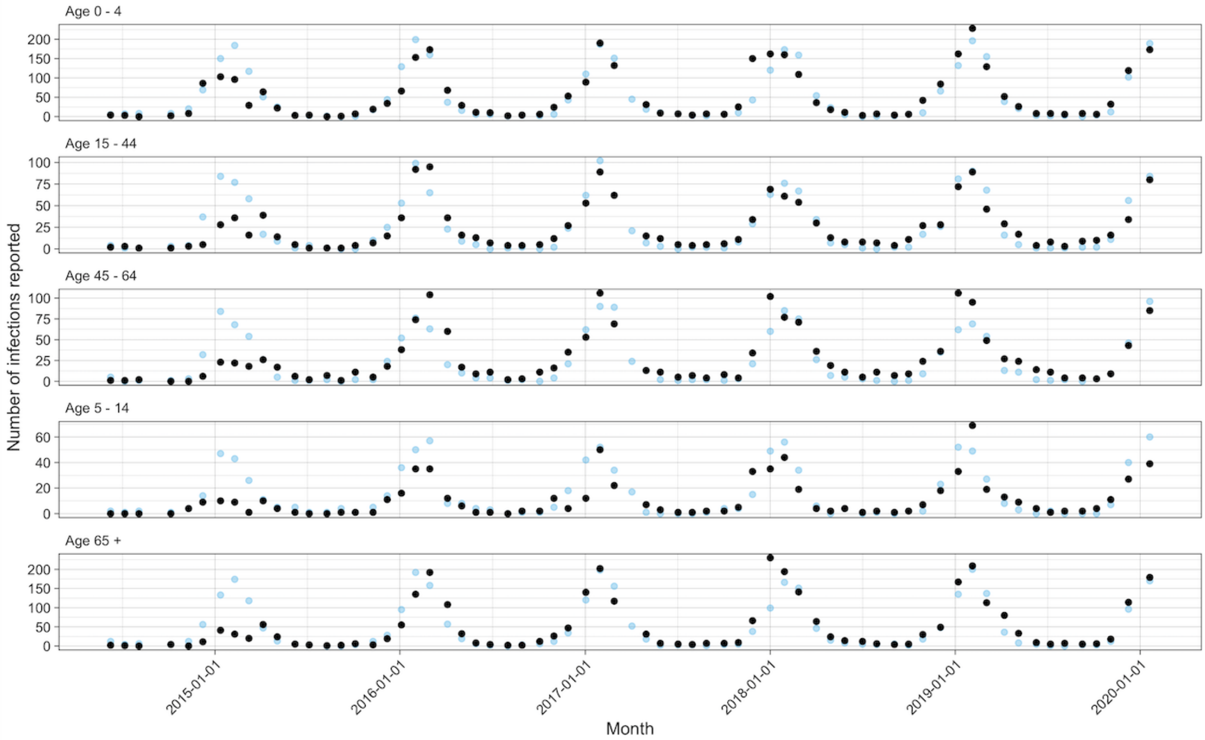


**Figure S7: Model fit for seasonal HCoV, using the immunity and $R_0$ parameters from Kissler et al. 2020.**

### 8. Additional projections

Figures S8 - S15 show additional projections for the next 30 years using different samples. The samples are using the same SARS-CoV-2 parameters, but with a different sample from the posterior estimates of seasonal coronavirus parameters.

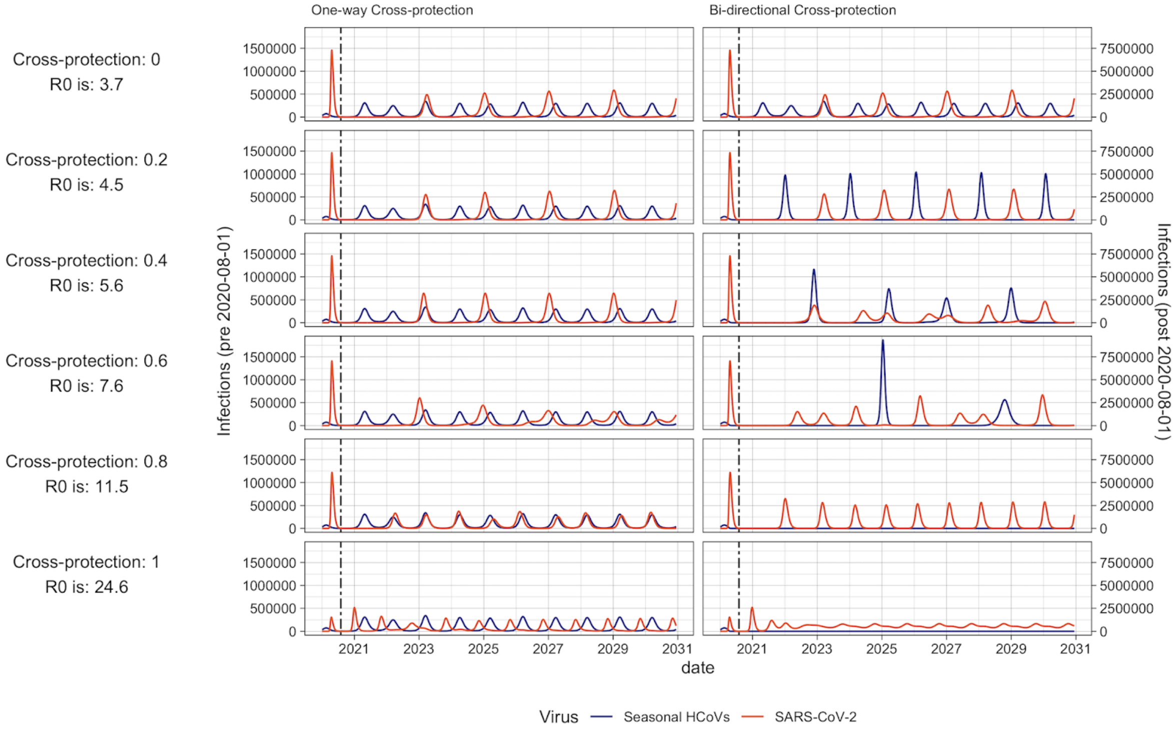


**Figure S8: Forward projections.** Projections of the number of daily infections for seasonal HCoVs and SARS-CoV-2 from the beginning of 2020 until the end of 2030. Red indicates SARS-CoV-2, blue indicates Seasonal HCoVs. The dashed vertical line indicates a change in scale, with the left hand side referring to the left y axis and right of the dashed line referring to the right y axis. Cross-protection strength and estimated $R_0$ for the scenario are shown to the left of the diagram, with descending rows indicating increased cross-protection. The left panel of the Figure depicts the projection with cross-protection only occurring from seasonal HCoV to SARS-CoV-2, whereas the right panel indicates bidirectional cross-protection. All simulations were run in the absence of any control measures.

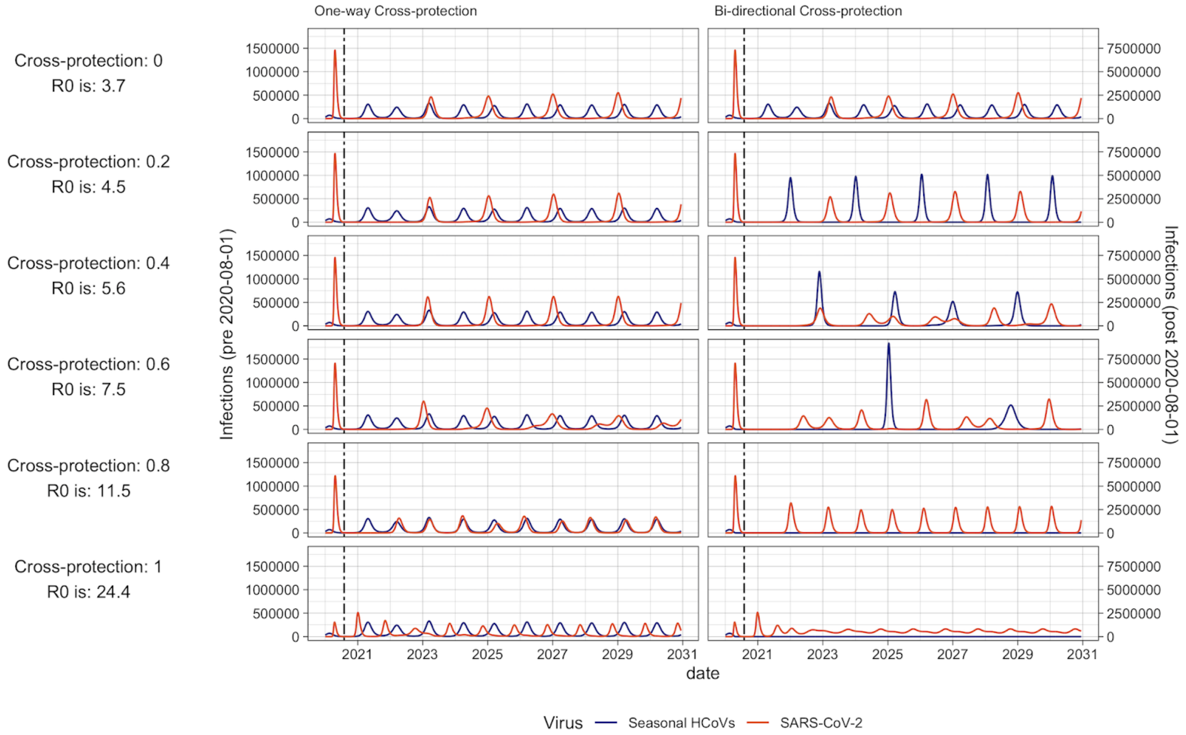


**Figure S9: Forward projections** Projections of the number of daily infections for seasonal HCoVs and SARS-CoV-2 from the beginning of 2020 until the end of 2030. Red indicates SARS-CoV-2, blue indicates Seasonal HCoVs. The dashed vertical line indicates a change in scale, with the left hand side referring to the left y axis and right of the dashed line referring to the right y axis. Cross-protection strength and estimated $R_0$ for the scenario are shown to the left of the diagram, with descending rows indicating increased cross-protection. The left panel of the Figure depicts the projection with cross-protection only occurring from seasonal HCoV to SARS-CoV-2, whereas the right panel indicates bidirectional cross-protection. All simulations were run in the absence of any control measures.

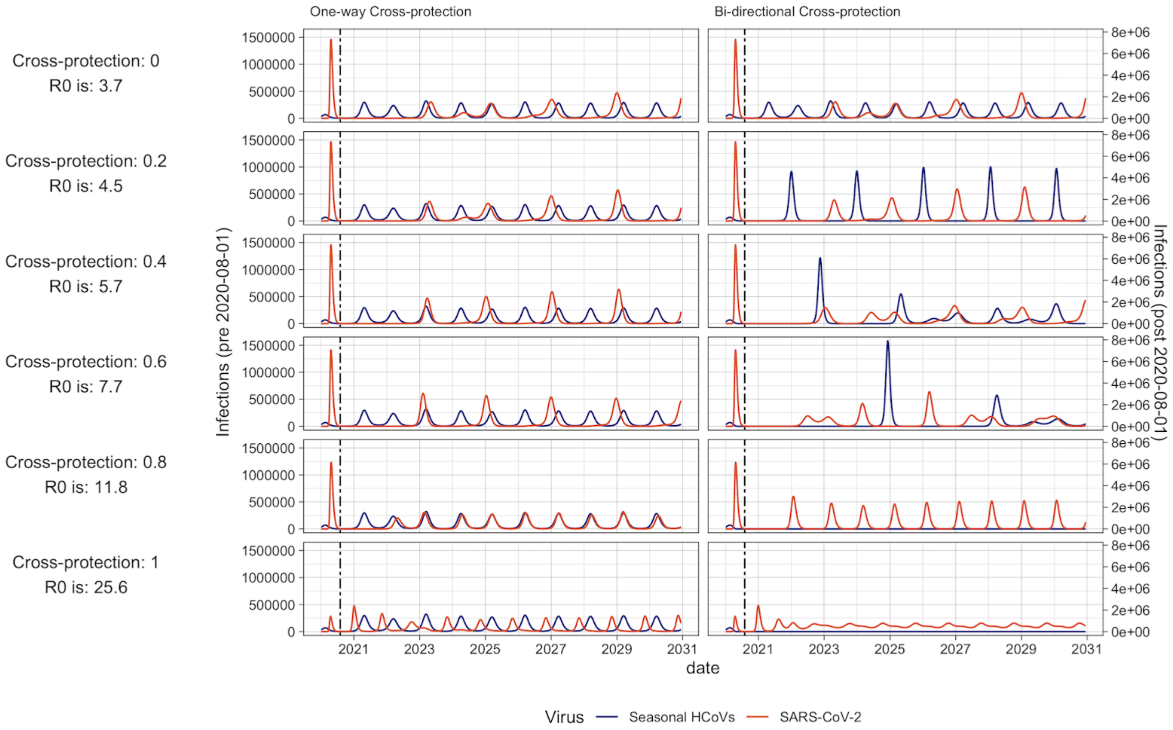


**Figure S10: Forward projections** Projections of the number of daily infections for seasonal HCoVs and SARS-CoV-2 from the beginning of 2020 until the end of 2030. Red indicates SARS-CoV-2, blue indicates Seasonal HCoVs. The dashed vertical line indicates a change in scale, with the left hand side referring to the left y axis and right of the dashed line referring to the right y axis. Cross-protection strength and estimated $R_0$ for the scenario are shown to the left of the diagram, with descending rows indicating increased cross-protection. The left panel of the Figure depicts the projection with cross-protection only occurring from seasonal HCoV to SARS-CoV-2, whereas the right panel indicates bidirectional cross-protection. All simulations were run in the absence of any control measures.

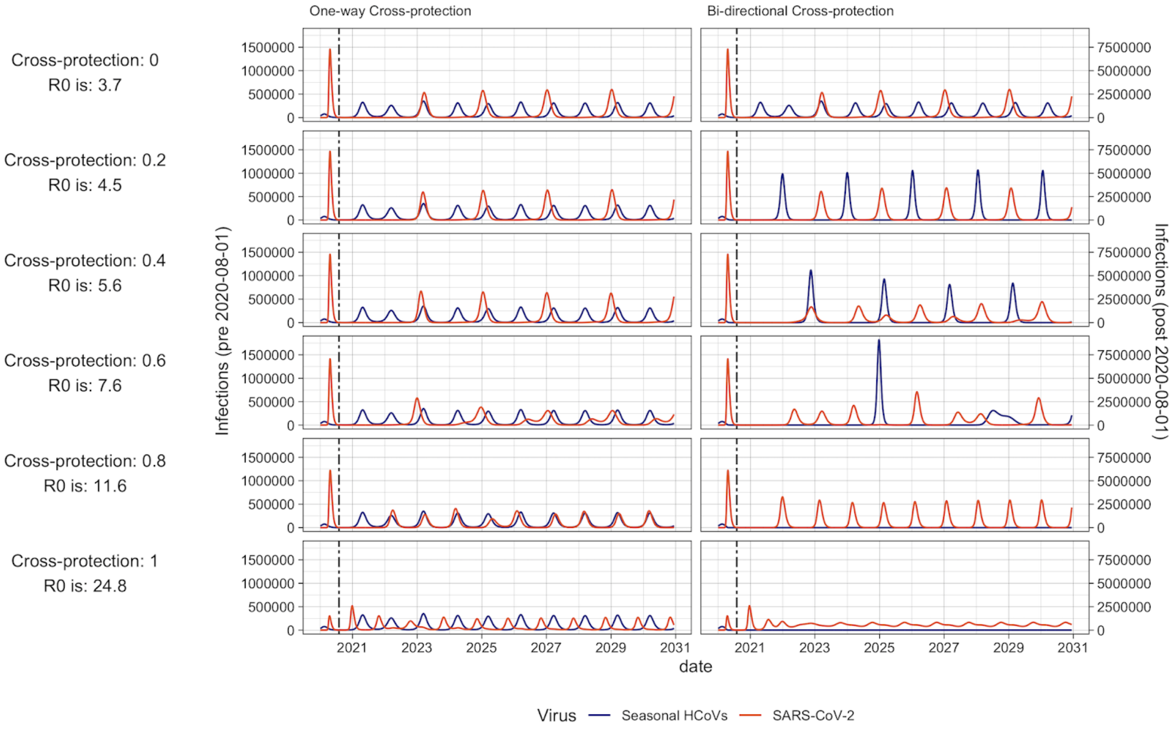

**Figure S11: Forward projections** Projections of the number of daily infections for seasonal HCoVs and SARS-CoV-2 from the beginning of 2020 until the end of 2030. Red indicates SARS-CoV-2, blue indicates Seasonal HCoVs. The dashed vertical line indicates a change in scale, with the left hand side referring to the left y axis and right of the dashed line referring to the right y axis. Cross-protection strength and estimated $R_0$ for the scenario are shown to the left of the diagram, with descending rows indicating increased cross-protection. The left panel of the Figure depicts the projection with cross-protection only occurring from seasonal HCoV to SARS-CoV-2, whereas the right panel indicates bidirectional cross-protection. All simulations were run in the absence of any control measures.

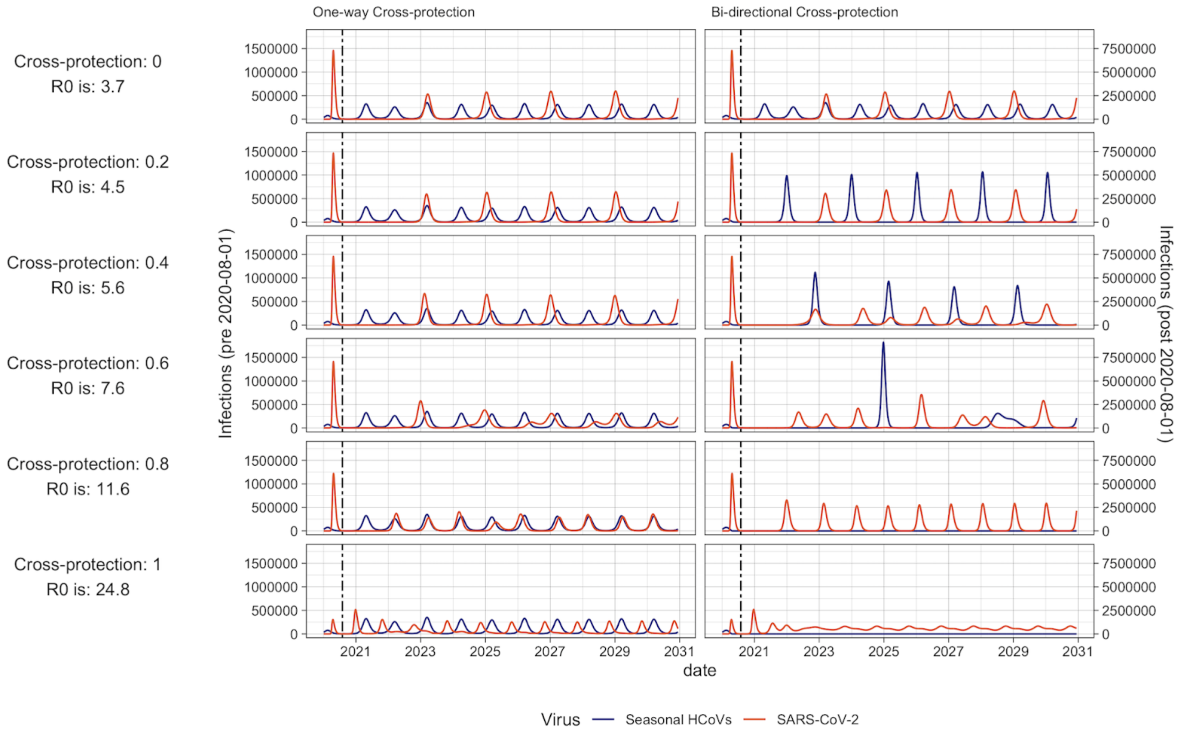


**Figure S12:Forward projections** Projections of the number of daily infections for seasonal HCoVs and SARS-CoV-2 from the beginning of 2020 until the end of 2030. Red indicates SARS-CoV-2, blue indicates Seasonal HCoVs. The dashed vertical line indicates a change in scale, with the left hand side referring to the left y axis and right of the dashed line referring to the right y axis. Cross-protection strength and estimated $R_0$ for the scenario are shown to the left of the diagram, with descending rows indicating increased cross-protection. The left panel of the Figure depicts the projection with cross-protection only occurring from seasonal HCoV to SARS-CoV-2, whereas the right panel indicates bidirectional cross-protection. All simulations were run in the absence of any control measures.

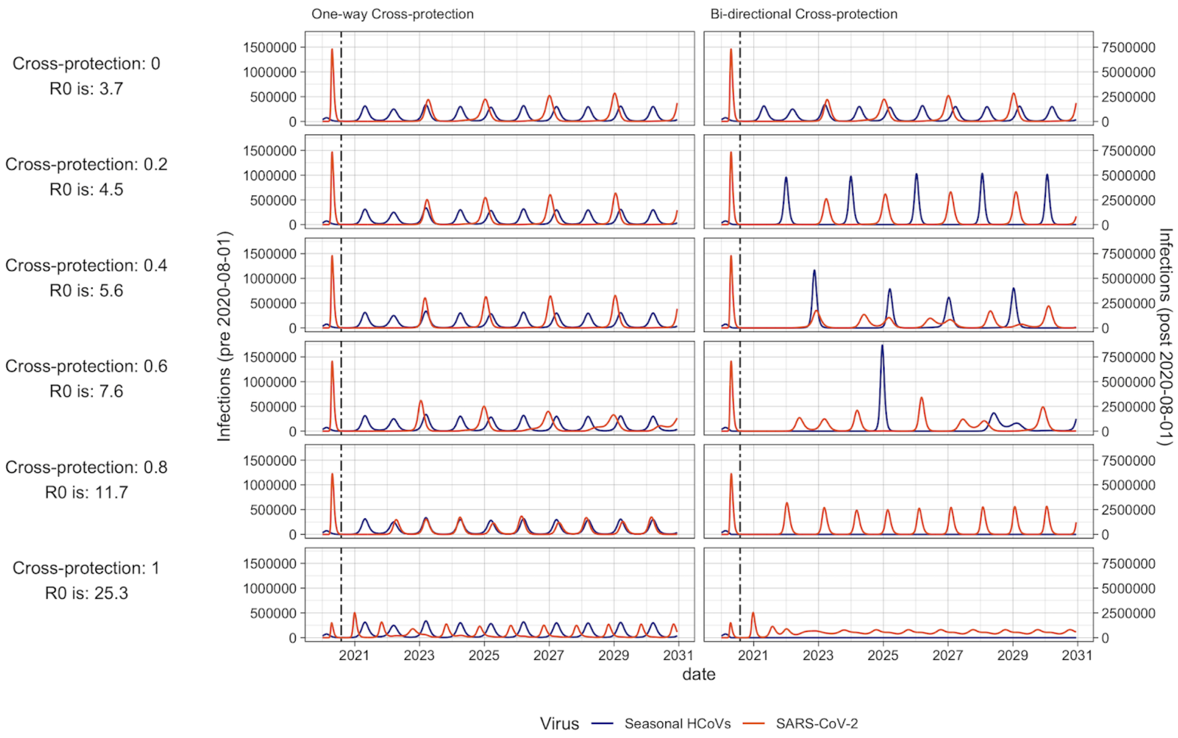


**Figure S13: Forward projections** Projections of the number of daily infections for seasonal HCoVs and SARS-CoV-2 from the beginning of 2020 until the end of 2030. Red indicates SARS-CoV-2, blue indicates Seasonal HCoVs. The dashed vertical line indicates a change in scale, with the left hand side referring to the left y axis and right of the dashed line referring to the right y axis. Cross-protection strength and estimated $R_0$ for the scenario are shown to the left of the diagram, with descending rows indicating increased cross-protection. The left panel of the Figure depicts the projection with cross-protection only occurring from seasonal HCoV to SARS-CoV-2, whereas the right panel indicates bidirectional cross-protection. All simulations were run in the absence of any control measures.

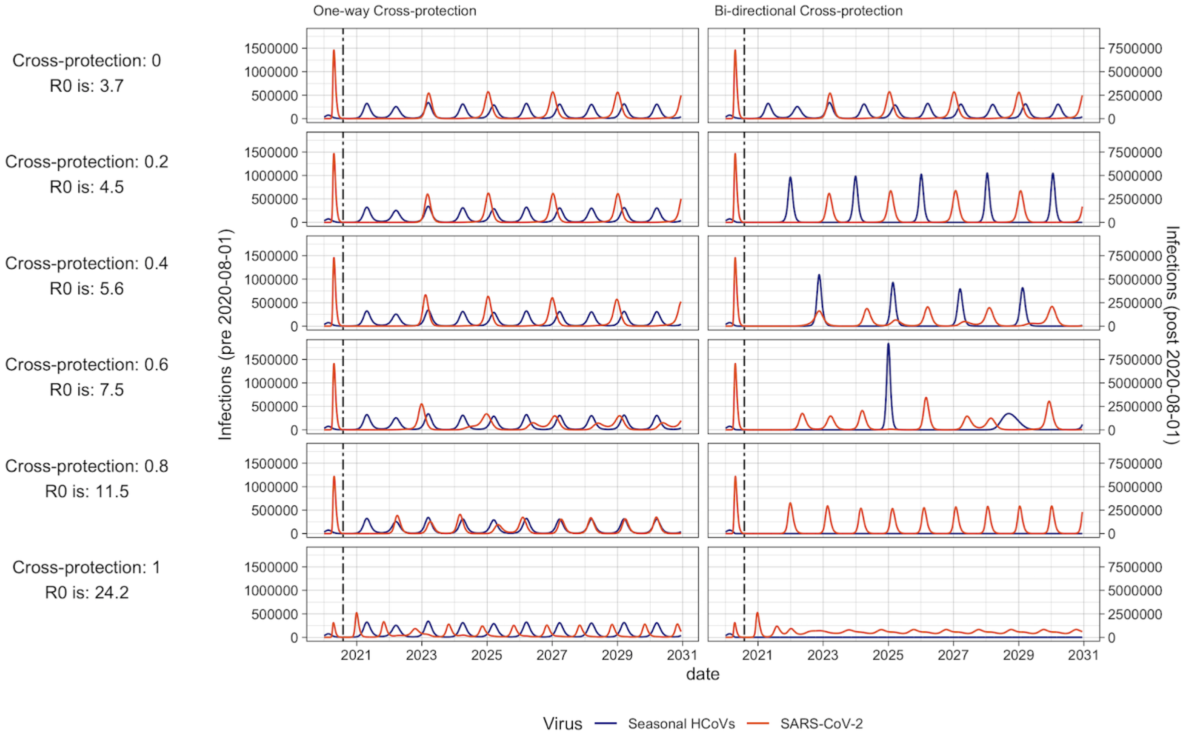


**Figure S14: Forward projections.** Projections of the number of daily infections for seasonal HCoVs and SARS-CoV-2 from the beginning of 2020 until the end of 2030. Red indicates SARS-CoV-2, blue indicates Seasonal HCoVs. The dashed vertical line indicates a change in scale, with the left hand side referring to the left y axis and right of the dashed line referring to the right y axis. Cross-protection strength and estimated $R_0$ for the scenario are shown to the left of the diagram, with descending rows indicating increased cross-protection. The left panel of the Figure depicts the projection with cross-protection only occurring from seasonal HCoV to SARS-CoV-2, whereas the right panel indicates bidirectional cross-protection. All simulations were run in the absence of any control measures.

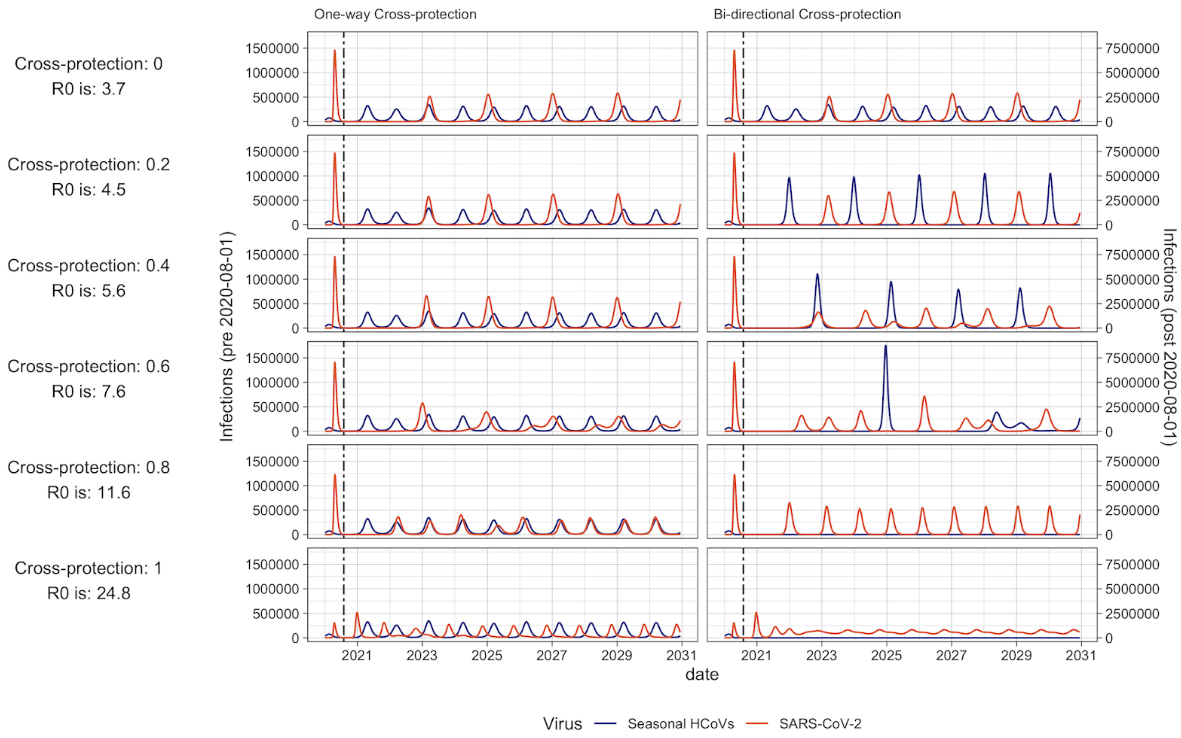


**Figure S15: Forward projections** Projections of the number of daily infections for seasonal HCoVs and SARS-CoV-2 from the beginning of 2020 until the end of 2030. Red indicates SARS-CoV-2, blue indicates Seasonal HCoVs. The dashed vertical line indicates a change in scale, with the left hand side referring to the left y axis and right of the dashed line referring to the right y axis. Cross-protection strength and estimated $R_0$ for the scenario are shown to the left of the diagram, with descending rows indicating increased cross-protection. The left panel of the Figure depicts the projection with cross-protection only occurring from seasonal HCoV to SARS-CoV-2, whereas the right panel indicates bidirectional cross-protection. All simulations were run in the absence of any control measures.

### 9. Seasonal sensitivity

We tested the sensitivity of our assumption of the seasonality period being 52 weeks, by running the model with 365.25 days in a year, using samples from our posterior. The resulting output is shown in Figure S14. We refit the seasonal timing ( $\varphi$ ) parameter using Maximum Likelihood, keeping all the other parameters constant at values from one sample of the posterior and calculated the likelihood values across the complete range of $R_0$ and duration of immunities. The resulting heatmap is shown in Figure S15, and our posterior estimate value is indicated with a “+”. This confirms that the estimated values of $R_0$ and waning duration are appropriate with the different assumptions.

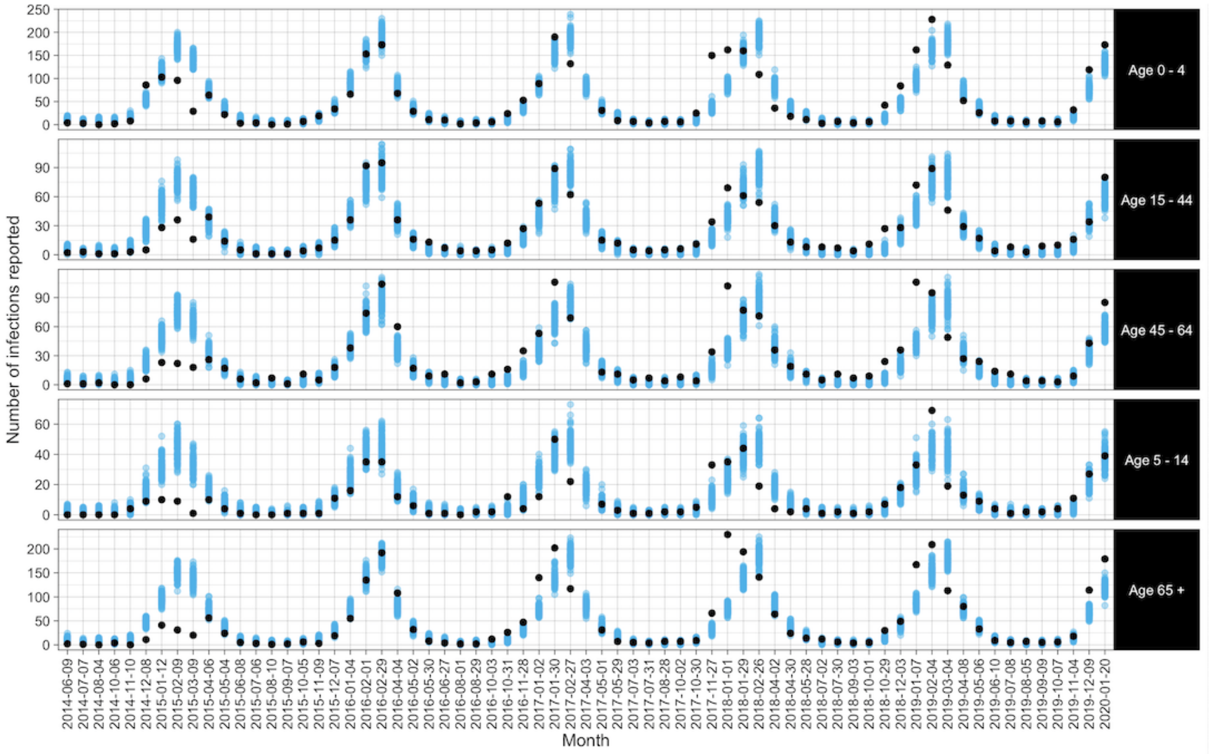


**Figure S16: Sensitivity analysis to seasonality function.** Fit to the data using our posterior parameter estimates and a model with a seasonality period of 365.25 days.

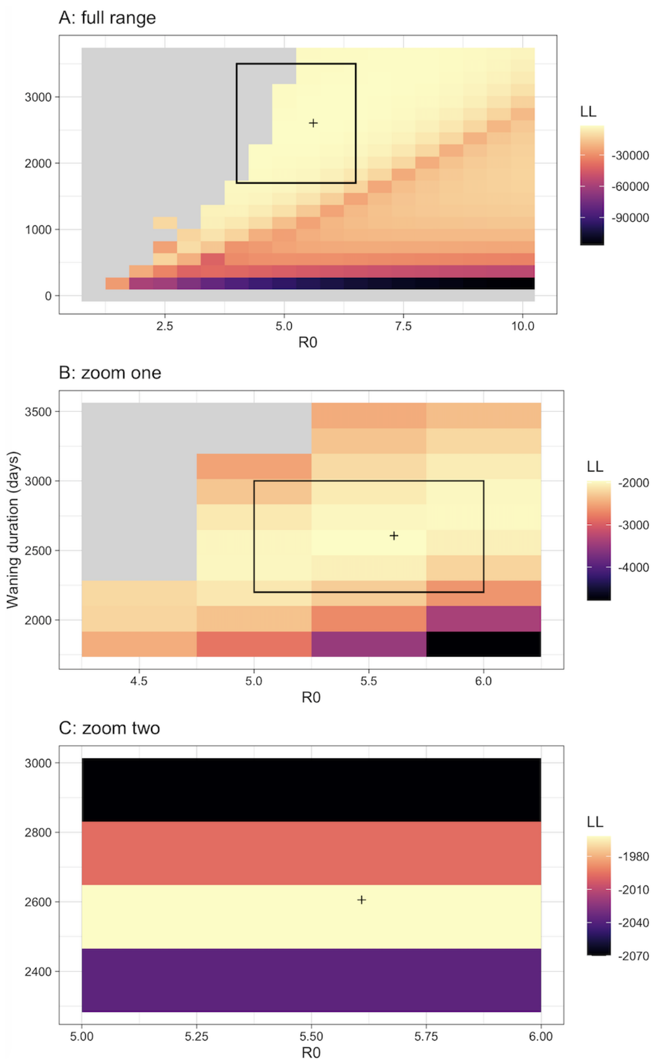


**Figure S17: Likelihood plane with varying $R_0$ and duration of waning, for the model with a seasonal period of 365.25 days.** A) shows the full range B) and C) show zoomed in areas. Colour indicates the likelihood value. The black rectangles show the area zoomed in on and the '+' symbol shows the estimated values from the parallel tempering fits.
